# RMS: A ML-based system for ICU Respiratory Monitoring and Resource Planning

**DOI:** 10.1101/2024.01.23.24301516

**Authors:** Matthias Hüser, Xinrui Lyu, Martin Faltys, Alizée Pace, David Berger, Marine Hoche, Stephanie L Hyland, Hugo Yèche, Manuel Burger, Tobias M Merz, Gunnar Rätsch

**Affiliations:** Department of Computer Science, ETH Zürich, Zürich, Switzerland; Swiss Institute for Bioinformatics, Lausanne, Switzerland; NEXUS Personalized Health Technologies, ETH Zürich, Zürich, Switzerland; Department of Intensive Care Medicine, University Hospital, University of Bern, Bern, Switzerland; Department of Intensive Care, Austin Hospital, Melbourne, Australia; AI Center, ETH Zürich, Switzerland; Microsoft Research, Cambridge, UK (current address); Cardiovascular Intensive Care Unit, Auckland City Hospital, Auckland, New Zealand; Medical Informatics Unit, Zürich University Hospital, Zürich, Switzerland; Department of Biology, ETH Zürich, Zürich, Switzerland

## Abstract

Acute hypoxemic respiratory failure (RF) occurs frequently in critically ill patients and is associated with substantial morbidity, mortality and increased resource use. We used machine learning to create a comprehensive monitoring system to assist intensive care unit (ICU) physicians in managing acute RF. The system encompasses early detection and ongoing monitoring of acute hypoxemic RF, assessment of readiness for tracheal extubation and prediction of the risk of extubation failure. In study patients, the model predicted 80% of RF events at a precision of 45%, with 65% of RF events identified more than 10 hours before RF onset. System predictive performance was significantly higher than standard clinical monitoring based on the patient’s oxygenation index and was successfully validated in an external cohort of ICU patients. We have demonstrated how the estimated risk of extubation failure (EF) could facilitate prevention of both, extubation failure and unnecessarily prolonged mechanical ventilation. Furthermore, we illustrated how machine-learning-based monitoring of RF risk, along with the necessity for mechanical ventilation and extubation readiness on a patient-by-patient basis, can facilitate resource planning for mechanical ventilation in the ICU. Specifically, our model predicted ICU-level ventilator use within 8 to 16 hours into the future, with a mean absolute error of 0.4 ventilators per 10 patients of effective ICU capacity.

## Introduction

Acute hypoxemic respiratory failure (RF) is a common occurrence in intensive care unit (ICU) patients and is associated with high morbidity, mortality and high resource use^1,2^. Hypoxemic RF (Type I RF) is the most common type of respiratory failure^3^ and its severity is defined by the P/F (PaO_2_/F_i_O_2_) ratio, with values below 200 mmHg corresponding to moderate and below 100 mmHg to severe RF. Treating patients with RF involves a sequence of clinical evaluations, including identifying RF and the need for mechanical ventilation, monitoring the recovery of lung function, determining the right time to stop mechanical ventilation, and assessing the risk of complications after tracheal extubation.

For optimal clinical decision-making, it is paramount to continuously monitor the patient’s clinical state in an attempt to predict their future clinical course. ICU physicians base their treatment decisions mostly on intermittent clinical assessments and trend evaluation of monitored vital signs stored in electronic patient data management systems. In the increasingly complex ICU environment, clinicians are confronted with large amounts of data from a multitude of monitoring systems of numerous patients. The quantity of data and the possibility of artifacts increases the risk that clinicians will not readily recognize, interpret, and act upon relevant information, potentially contributing to suboptimal patient outcomes and increased ICU resource expenditure^4^ compared to optimal care. Large datasets involving multiple data points on many patients are ideal for automatic processing by machine learning (ML) algorithms^5,6^. To facilitate such advancements, we previously published the High time Resolution Intensive care unit (HiRID) Dataset, which encompasses approximately 34,000 ICU admissions^12^. ML has been used to develop decision support systems for various conditions in the ICU, such as acute respiratory distress syndrome (ARDS)^7–11^, circulatory failure^12^, sepsis^13–15^, and renal failure^16^.

We aimed to develop a comprehensive, ML-based respiratory monitoring system (RMS), consisting of multiple subsystems to simplify and expedite the management of individual patients with RF and to optimize ICU resource planning. For individual patients, the system predicts the risk of hypoxemic RF (RMS-RF) and the need for mechanical ventilation (RMS-MV_Start_), continuously monitors changes and improvements of the respiratory state, and predicts the remaining time of required mechanical ventilation (RMS-MV_End_) and probability of successful extubation (RMS-EF). We investigated how using respiratory state predictions on a patient-by-patient basis could enable estimating the future number of patients in need of mechanical ventilation on a shift-to-shift short-term basis (resource planning). In addition, we prepare a new version of the dataset, HiRID-II, which we anticipate will significantly expand both the number of included patients and the range of available clinical variables.

We hypothesized that our ML-system could predict the relevant respiratory events throughout the treatment process of individual patients accurately and early; both in the development dataset and when validated in externally sourced data. In addition, we intended to develop a resource management support tool to predict ICU-level future mechanical ventilator use by integrating all RMS scores across ICU patients.

## Results

### Preparation of an extended HiRID dataset (HiRID-II)

We present the High time Resolution Intensive care unit Dataset II (HiRID-II), a substantial update to HiRID-I^17^, that will be made available^1^ to the research community on Physionet.org^18,19^. This new dataset contains 60% more ICU admissions than its predecessor (**Table 1, Extended Data Fig. 1a**). Additionally, the number of variables has increased from 209 to 310 (**Extended Data Fig. 1b**). The dataset was k-anonymized in regard to age, weight, height, and gender, reducing the number of admissions from 60,503 to 55,858^20^. Admission dates were randomly shifted to further reduce the risk of identification of individual patients. To allow the assessment of model generalization to the future, the dataset was divided into temporal splits while respecting k-anonymization (**Extended Data Fig. 1c**). A high-resolution external evaluation dataset, extracted from the Amsterdam UMCdb^21^, was used to test the generalizability of RMS to other health care systems (**Extended Data Fig. 1d**). Preliminary analysis of the HiRiD-II dataset revealed strong correlations between the occurrence of RF and EF with ICU mortality, confirming prior results^1^ and motivating our proposed RMS (**Extended Data Fig. 2**).

**Table 1:**
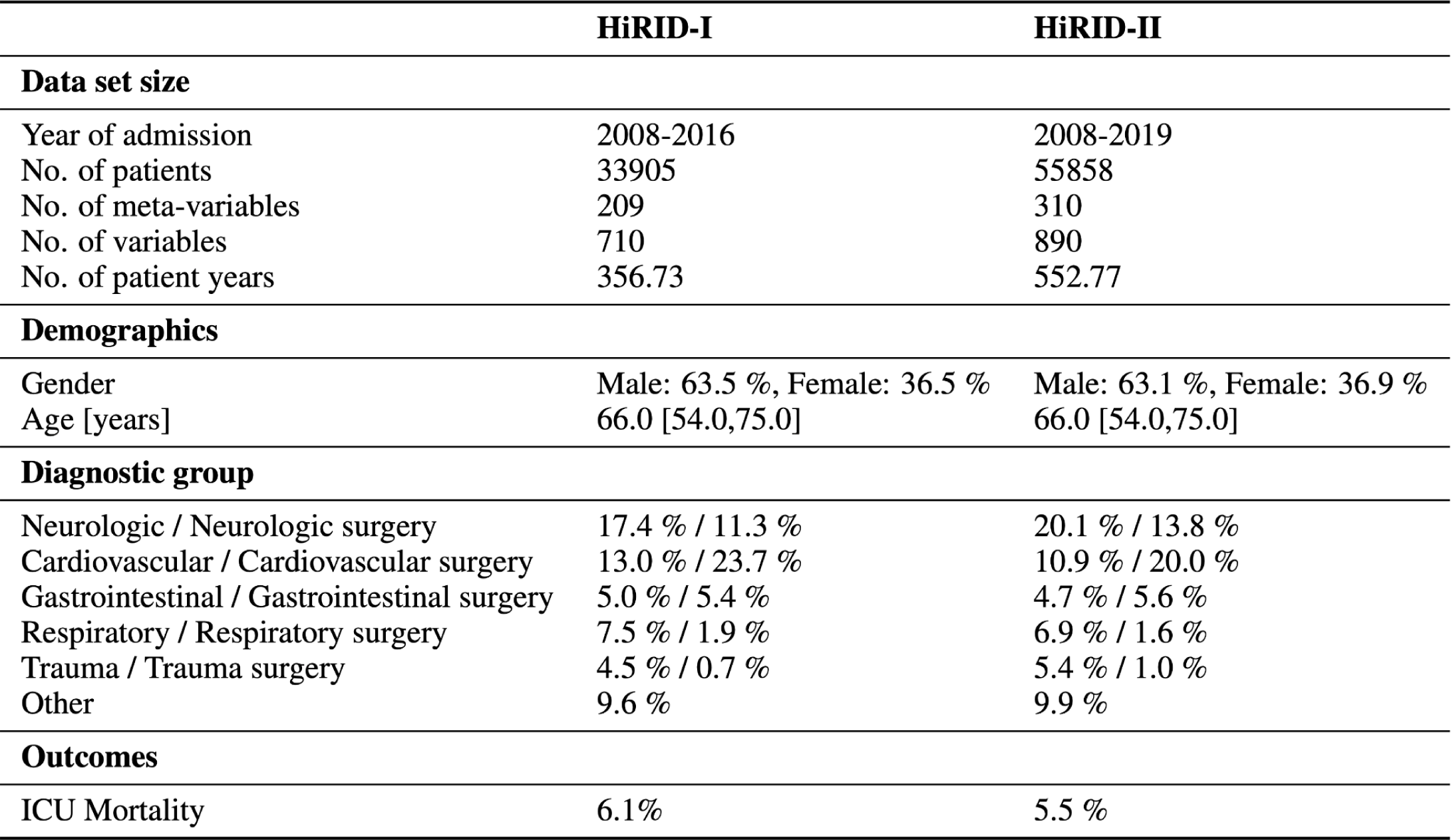
Characteristics of the HiRID-I and HiRID-II datasets. Age is reported as median and interquartile range (IQR). The statistics were computed on the HiRID datasets after k-anonymization. HiRID-II includes also patients from HiRID-I.

### Development of a continuous monitoring system for respiratory management

We developed a comprehensive ML-based respiratory monitoring system composed of four interrelated predictive models that, together, cover the respiratory trajectory of patients. RMS-RF estimates every five minutes the risk of moderate to severe hypoxemic respiratory failure (P/F ratio < 200mmHg) in the next 24 hours. RMS-MV_Start_ predicts the need for mechanical ventilation, while RMS-MV_End_ determines if the patients will be ready to be liberated from ventilatory support; both tasks forecast risk within the subsequent 24 hours. Finally, RMS-EF evaluates the likelihood of successful extubation at given time points, when the patient already meets formal criteria.

#### Patient state annotation and labeling

For each time point we determined if a patient is currently in moderate or severe hypoxemic RF (P/F ratio < 200 mmHg), mechanically ventilated, or ready to be extubated. Readiness to extubate (REXT) status at each time point was defined using a heuristic scoring system determined by gas exchange, respiratory mechanics, hemodynamics and neurological status. A score threshold was manually selected after inspection of the time series by an experienced ICU clinician (**Fig. 1b**). At these time-points the patient could be extubated according to formal criteria, but extubation failure can still occur. The current ventilation status was derived from the presence of ventilator-specific parameters.

**Fig. 1:**
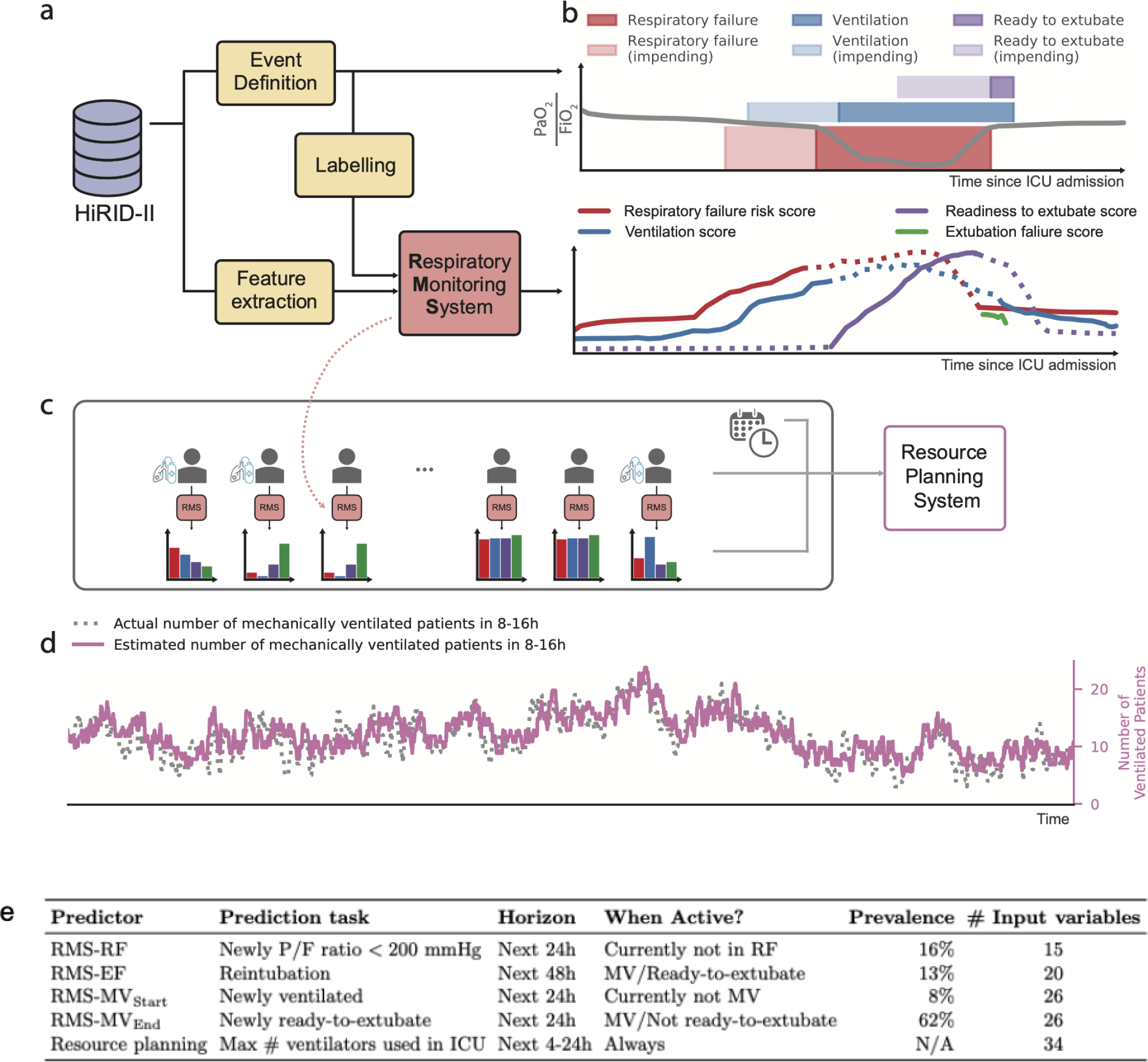
Overview of the RMS decision support system for Respiratory State Management, and its extension for ICU-level resource planning. **a.** Flow diagram for the development of RMS predictors at the individual patient level. Time series were extracted from the HiRID-II dataset and gridded to a five-minute resolution, and features were computed. Respiratory failure/ventilation/ready-to-extubate periods were annotated and machine learning labels created. **b.** The respiratory monitoring system consists of four scores which are active during different time periods of the ICU stay, according to the current respiratory and ventilation state of the patient **c.** Flow diagram for the development of a resource monitoring system at the ICU level. For all current patients in the ICU, the four scores were integrated to predict the probability that a patient will require mechanical ventilation within a future time horizon. **d.** Example time period of 3 months, displaying the actual number of ventilated patients and the predicted number as estimated by RMS in the next 8 to 16 hours. **e.** Overview of prediction tasks solved by RMS for individual patients (RMS-RF/RMS-EF/RMS-MV_Start_/RMS-MV_End_) as well as on the ICU-level. For RF, MV_Start_ and MV_End_ we provide the event prevalences in the test set at times when the patient is stable, not ventilated, or ventilated, respectively.

Positive labels for future RF were defined as time points when the patient was not currently in RF but would exhibit RF in the next 24 hours (“impending RF”); while negative labels were assigned to time points when the patient was not currently in RF and would remain stable during the next 24 hours. For every extubation event, we determined whether it failed (reintubation necessary within 48h after extubation) and used it as the label for extubation failure. Labels for ventilation onset and readiness to extubate prediction were positive at time points when the patient was currently not ventilated/ready-to-extubate but would be in the next 24 hours (**Fig. 1b**). In HiRID-II, 43.7% and 46.2% of all patients had RF events and required mechanical ventilation, respectively. The dataset contained 23,861 extubation events, of which 11.1% extubations failed.

#### Continuous P/F ratio estimation

To measure PaO_2_, an arterial blood sample is necessary. PaO_2_ is therefore only periodically available at a low temporal resolution. For continuous high-resolution hypoxemic RF labelling, a continuous estimation of the current PaO_2_ is required. An underlying physiological principle determines the binding and dissociation of oxygen and hemoglobin, yielding a sigmoid correlation between arterial oxygen saturation (SpO_2_) and PaO_2_ ^24–26^. A model based on the continuously available SpO_2_ can therefore estimate continuous PaO_2_. We developed an ML algorithm to produce continuous estimates of PaO_2_ based on SpO_2_ and other relevant variables determining the hemoglobin-oxygen dissociation curve. The algorithm outperformed the existing non-linear Severinghaus-Ellis baseline^27^ for estimating PaO_2_ values from non-invasive SpO_2_ measurements (**Extended Data Fig. 3**). By integrating PaO_2_ estimates with continuously available FiO_2_ data, we derived P/F ratio estimates on a five-minute time grid.

#### Development of RMS predictors

The RMS produces four individual scores active at different stages of the RF management process. All four subsystems are based on manual feature engineering and LightGBM^22^ predictors, similar to our previous work^12^. Prior analyses on HiRID-I for circulatory failure and a related respiratory failure task have shown superior performance of LightGBM compared to other models including deep learning^12,23^. The predictor for RF (RMS-RF) used 15 clinical variables (**Supplemental Table 3)**. As in Hyland et al.^12^, the system triggered an alarm if the RF score exceeded a specified threshold. It was silenced for 4 hours afterwards. The alarm system was reset when the patient recovered from an event and could reactivate 30 minutes later. The extubation failure (RMS-EF) predictor used 20 clinical variables (**Supplemental Table 3)**. The RMS-RF & RMS-EF variable sets were identified separately for the two tasks, using greedy forward selection on the validation set of five data splits. The models for ventilator use (RMS-MV_Start_) and extubation readiness (RMS-MV_End_) used the union of the parameters of the two main tasks, consisting of 26 variables in total (**Supplemental Table 3**).

We utilized the four risk scores to predict short-term mechanical ventilator resource requirements by training a meta-model (**Fig. 1c**). The resource planning problem was divided into two sub-problems; predicting the future ventilator use for already admitted ICU patients, and predicting the requirement for mechanical ventilation for newly admitted non-elective patients in the near future. We excluded elective patient admissions as their resource use is typically known well in advance (hence, no prediction is needed). The predictor uses date and time information as well as summary statistics regarding ventilator use and patient numbers derived from the ICU dataset. A LightGBM^22^ regressor was trained to solve both sub-problems. For admitted ICU patients, it predicts the necessity for mechanical ventilation in the short-term future, as well as the total number of ventilators required for all admitted patients as an aggregate of the individual predictions (**Fig. 1d**).

#### Open source release

All elements of the developed system (**Fig. 1e**), including data preprocessing, annotation, prediction task labeling, and both training and prediction pipelines are made available under an open source license facilitating the reproducibility and reuse of the methodology and results.

### RMS-RF predicted RF early with high precision and reduced false alarms compared to clinical baselines

We developed a model that continuously evaluates the likelihood of a patient developing hypoxemic RF within the next 24 hours, updating its predictions every five minutes throughout the ICU stay. We define RF as a moderate or severe reduction in oxygenation, reflected by a P/F ratio below 200 mmHg. To focus on impending deterioration, the model only generates predictions when the patient is not currently experiencing RF. Conversely, if the patient remains stable and does not meet criteria for RF over the subsequent 24 hours, the model recognizes a low-risk state. The early prediction of hypoxemic RF is crucial for timely intervention and may reduce the number of unfavorable patient outcomes and improve overall healthcare quality. By accurately forecasting these events, RMS-RF may not only improve clinical decision-making but also allow physicians to commence treatment early, thereby mitigating the risk of more severe respiratory complications.

The RMS-RF model achieved an area under the alarm/event precision recall curve^12^ (AUPRC) of 0.559 with an alarm precision of 45% at an event recall of 80%. The model had an area under the receiver operating characteristic curve (AUROC) of 0.839 (**Extended Data Fig. 4a**). We observed that RMS-RF significantly outperformed two comparator baselines, a decision tree that uses the current value of respiratory and other parameters (SpO_2_, F_i_O_2_, PaO_2,_ Positive end-expiratory pressure (PEEP), RR, Ventilator presence, HR, GCS) as well as a clinical threshold-based system based on the SpO_2_/F_i_O_2_ ratio (**Fig. 2a**). RMS-RF was well calibrated, in contrast to the two baselines (**Extended Data Fig. 4b**). The system detected 65% and 78% of respiratory failure events at least 10 hours before they occurred when set to an event recall of 80% and 90% respectively (**Fig. 2b**). Compared to the SpO2/FiO2 threshold, our system generated two-thirds fewer false alarms per day on days without respiratory failure. (**Fig. 2c**). The model performance increased with inclusion of data up to 25% of the total dataset size, while no further improvements were observed when using more data (**Extended Data Fig. 4c**). Model performance was highest in patients across cardiovascular and respiratory diagnostic groups (alarm precisions 55% and 60% at 80% event recall, respectively). Lower performance was observed in neurologic and trauma patients (**Fig. 2d**). Performance varied in groups determined by age and gender^28^ (**Extended Data Fig. 4d/e**). RMS-RF exhibited physiologically plausible relationships between risk and clinical variables, according to SHapley Additive exPlanations (SHAP)^29^ values (**Fig. 2e**, **Extended Data Fig. 5**).

**Fig. 2:**
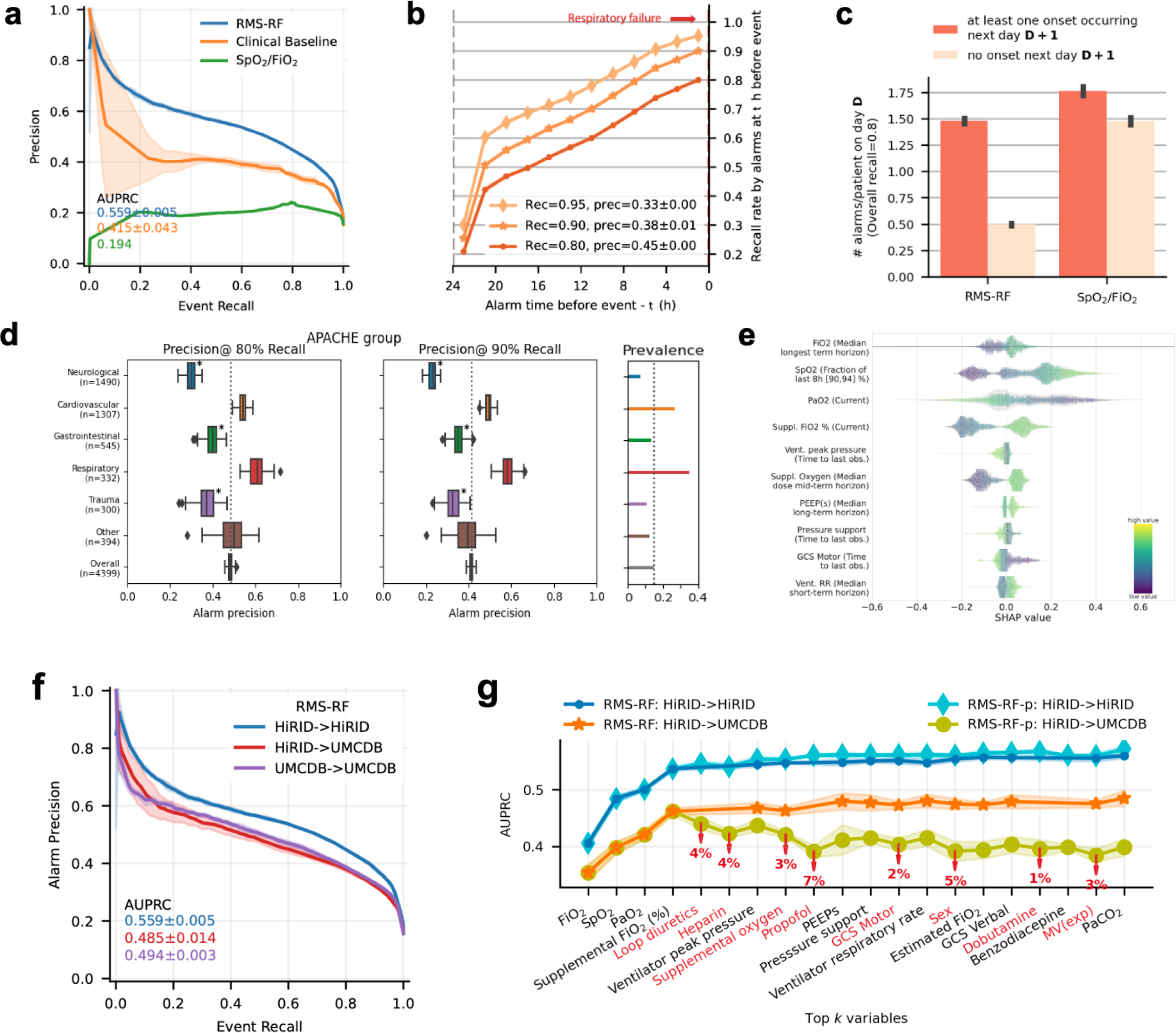
RMS-RF: Model performance / feature inspection of the respiratory failure prediction model. **a.** Model performance of RMS-RF compared with a decision-tree based clinical baseline and a threshold-based alarm system based on the current SpO_2_/F_i_O_2_ ratio. **b.** The RMS-RF system’s performance was evaluated in terms of earliness of its alarms. Specifically, this is measured as the proportion of respiratory failure events for which it provides early warning during a fixed prior time period and considering only events with a prior sufficient stability period. **c.** Comparison of generated false/true alarm counts of RMS-RF compared with a fixed threshold alarm system, both for patients with events, and patients without events on a given day. **d.** Alarm precision at event recalls of 80% and 90% of the RMS-RF model by admission diagnostic group category. The model was re-calibrated for each sub-group using information available at admission time, to achieve a comparable event recall. **e.** Feature inspection using SHAP values for the most important features for predicting respiratory failure, depicting the relationship between feature values and SHAP values. **f.** External validation of RMS-RF in the Amsterdam UMCdb dataset^21^. Internal, transfer as well as retrain performance in UMCdb is displayed. **g.** RMS-RF performance changes as the most important variables are added incrementally to the model, for the internal HiRID setting, and the transfer setting to UMCdb. Model transfer issues between the two hospital centers existed if medication variables were included in RMS-RF, denoted as the RMS-RF-p model variant. Markers denote the variables included in the models, and red colors denote variables which decrease performance when added to the model in the transfer setting.

The proposed RMS-RF model used a small number of physiological parameters and ventilator settings. Slightly diminished performance was observed when the HiRID-II-based model was externally validated in the Amsterdam UMCdb dataset^21^. There was no significant performance improvement observed through retraining with local data (**Fig. 2f**; both have 38% alarm precision at 80% event recall). We excluded medication variables to reduce the effect of differences in medication policies in different hospitals. A variant of RMS-RF including medication variables (RFS-RF-p) achieved only minor gains in internal HiRID performance (**Fig. 2g**) and exhibited poor transfer performance to UMCdb (**Extended Data Fig. 6a**). Medication policy differences between HiRID-II and UMCdb were analyzed to investigate these drops in transfer performance, indicating that these were generated by differences in the use of loop diuretics, heparin, and propofol (**Fig. 2g**, **Extended Data Fig. 6b/c**).

### RMS-EF predicted extubation failure with high precision and was well-calibrated

The accurate prediction of extubation failure is a critical aspect of patient management in intensive care, enabling clinicians to make informed decisions about the ideal timing of extubation. By utilizing RMS-EF to predict the risk of extubation failure, physicians could judiciously determine whether to proceed with or delay extubation based on a quantifiable risk threshold, potentially reducing the likelihood of complications associated with both, premature extubation or unnecessary prolongation of mechanical ventilation. We compared the developed RMS-EF predictor to a threshold-based scoring system, which counts the number of violations of clinically established criteria for readiness to extubate at the time point when the prediction is made (REXT status score). RMS-EF significantly outperformed the baseline (**Fig. 3a**) with an AUPRC of 0.535 and an AUROC of 0.865 (**Extended Data Fig. 7a**). We also analyzed calibration and observed a high concordance between observed risk of extubation failure and RMS-EF with a Brier score of 0.078; in contrast to the baseline (**Fig. 3b**). The precision for predicting extubation failure was 80% at a recall of 20% indicating that RMS-EF can effectively identify the patients at highest risk. RMS-EF predicted successful extubation at least 3h prior to the time point when extubation effectively takes place in 25% of events (**Fig. 3c**). As with RMS-RF, no major improvements in model performance were observed when using more than 25% of the training data (**Extended Data Fig. 7b**). Performance across diagnostic groups was similar, with RMS-EF performing best in respiratory patients (**Fig. 3d**). We observed that the performance in female patients and older age groups was slightly inferior (**Extended Data Fig. 7c/d**). As RMS-EF is based almost exclusively on variables that are influenced by clinical policies (**Fig. 3f**, **Extended Data Fig. 8**) which likely differ in different hospitals, it transferred poorly to the UMCdb dataset^21^ (**External Data Fig. 7e**). However, a variant of our model can be constructed without medication variables, which transferred well to the UMCdb dataset with slightly reduced internal performance (**Fig. 3e**; AUPRC 53.5% vs. 49% for HiRID). Accordingly, the analysis of medication policies revealed major differences for ready-to-extubate patients between HiRID-II and UMCdb (**Extended Data Fig. 7f/g**). SHAP value analysis^30^ showed that the RMS-EF risk score was dependent on several parameters determined by treatment-policies (**Fig. 3f**, **Extended Data Fig. 8**). Severe loss of transfer performance resulted from the inclusion of sedatives and vasopressors in the model (**Fig. 3g**).

**Fig. 3:**
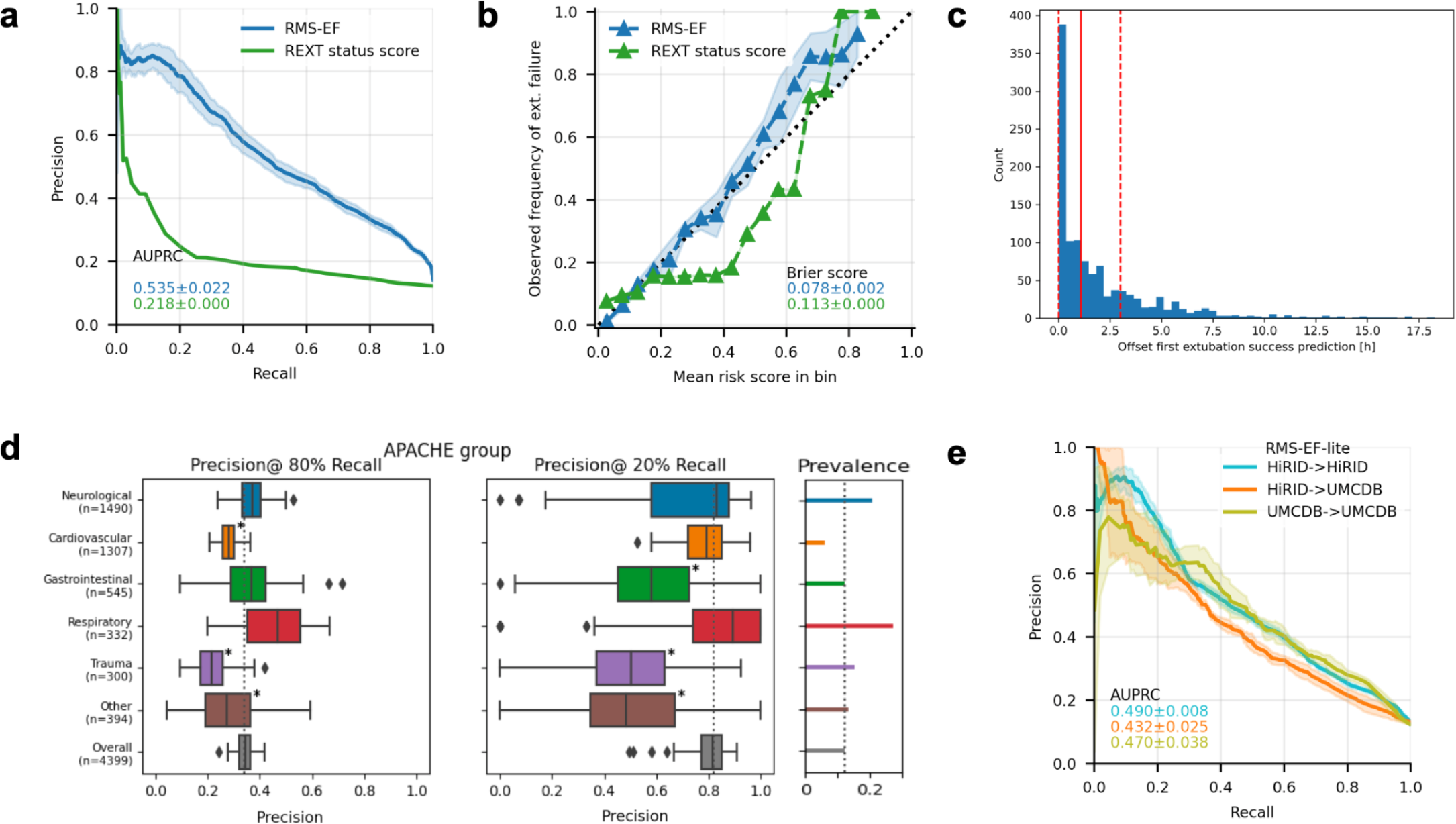
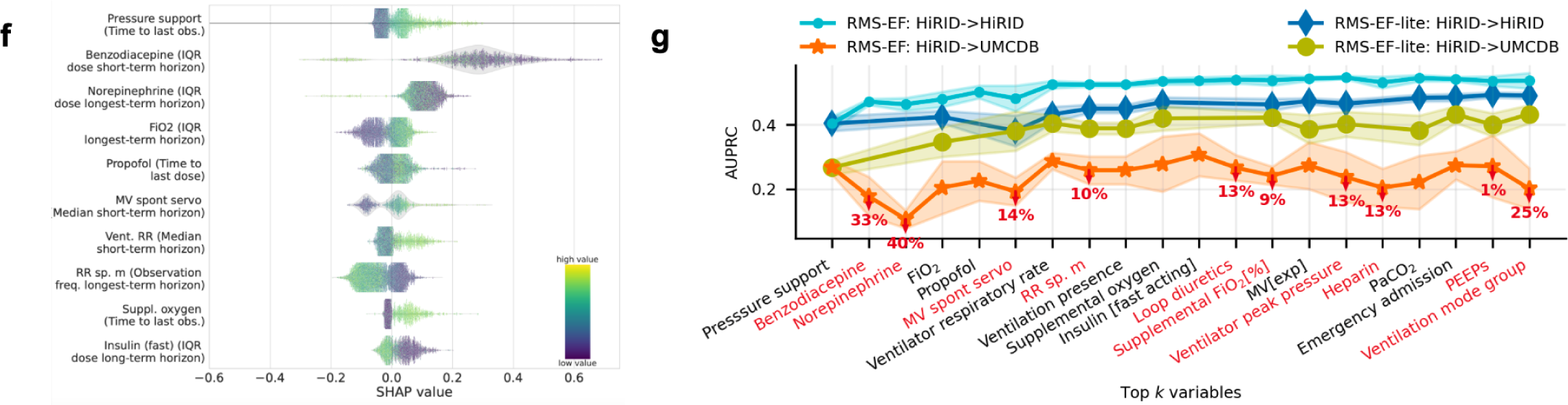
RMS-EF: Model performance analysis and feature inspection. **a.** Model performance compared with a baseline based on clinically established criteria for readiness to extubate in terms of recall/precision. **b.** Risk calibration of the score for predicting extubation failure at the time of extubation, compared with the baseline. **c.** Distribution of time span between the earliest extubation success prediction of RMS-EF prior to the time point of successful extubation, for correctly predicted successful extubations. The earliest time is defined as the first time point from which RMS-EF continuously predicts ‘extubation success’ while the patient is ready-to-extubate. Red dashed lines denote the 25, 75 percentiles, and the red solid line denotes the median, respectively. **d.** Performance stratified by admission diagnostic group in terms of precision at 80% and 20% recall. The model was re-calibrated for each sub-group using information available at admission time, to achieve a comparable recall. A * next to a bar indicates significantly different from average performance. **e.** Performance of the RMS-EF-lite model, which is obtained by excluding medication variables from RMS-EF, when trained/tested on the HiRID-II dataset, transferred to the UMCdb dataset, and retrained in the UMCdb dataset. **f.** Summary of SHAP value vs. variable distribution for the most important feature of each of the top 10 important variables contained in the RMS-EF model. **g.** Performance of the RMS-EF and RMS-EF-lite models in the internal and transfer settings as variables are added incrementally to the model ordered by performance contribution (greedy forward selection performance on the validation set). Red marked percentages on the orange curve denote relative performance loss in the transfer, when adding the variable to the model. Variables are in red font if their inclusions lead to performance loss in the transfer setting.

### Predicting intubation and readiness-to-extubate for individual patients

We evaluated prediction of ventilation onset (RMS-MV_Start_) and readiness to extubate (RMS-MV_End_) within the next 24h also on a patient-level. We observed high discriminative performance with AUROCs of 0.914 and 0.809 (**Extended Data Fig. 9a/b**), event-based AUPRCs of 0.528 and 0.910 (**Extended Data Fig. 9c/d**), for RMS-MV_Start_ and RMS-MV_End_ respectively, and the models were well calibrated (**Extended Data Fig. 9e/f**).

### Integrating all RMS scores of individual patients for planning ICU-level resource allocation

Using the predictions for the four models focusing on respiratory failure (RMS-RF), extubation failure (RMS-EF), ventilation onset (RMS-MV_Start_), and readiness to extubate (RMS-MV_End_), we developed a combined model predicting the number of ventilators in use for non-elective patients at a specific future horizon. Preliminary analysis of the HiRID-II dataset demonstrated substantial variation in demand for ventilators each day, underscoring the need for a model to aid resource planning (**Fig. 4a**).

**Fig. 4:**
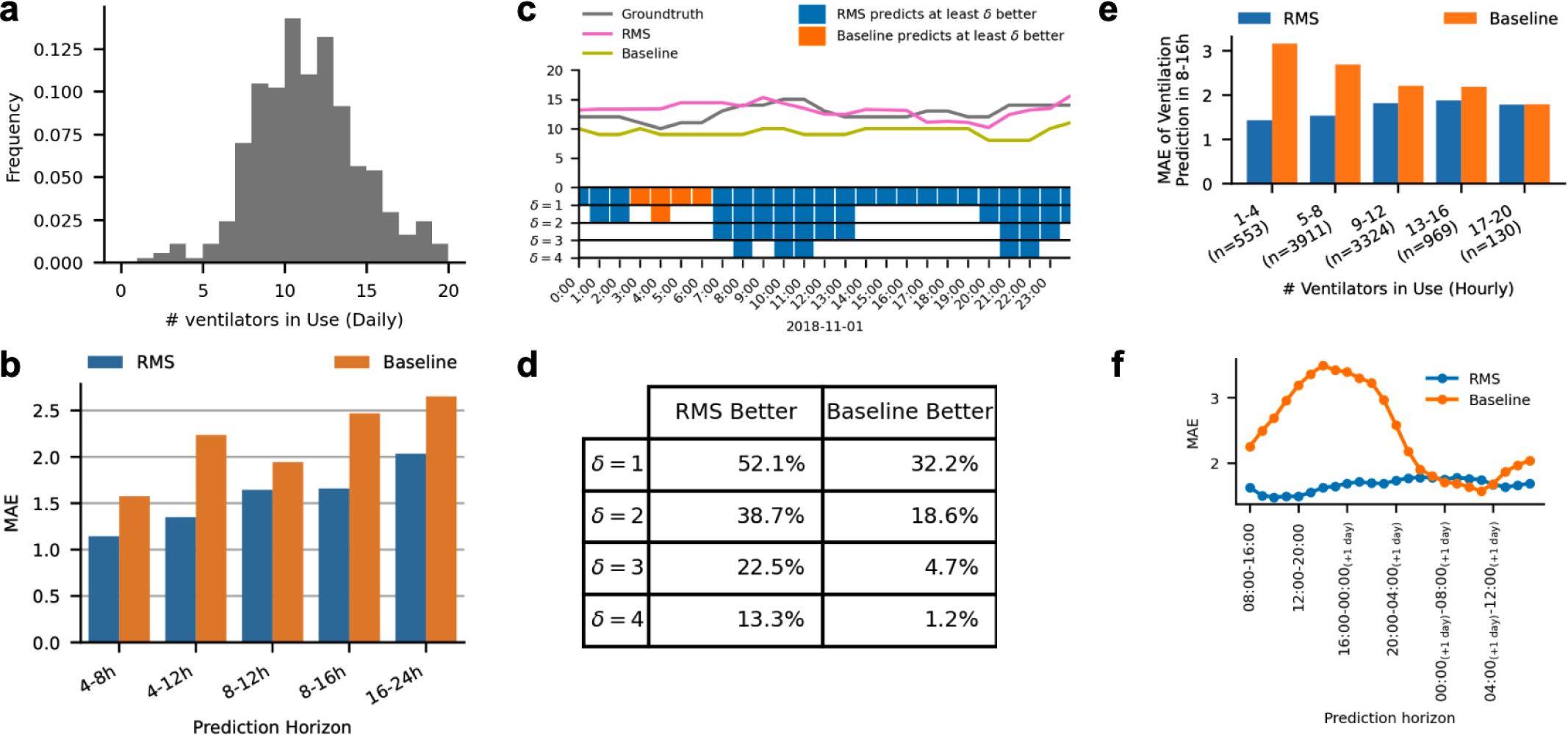
Model performance of the integrated system for resource planning (RMS) in an ICU with an effective capacity of 42 beds. **a.** Observed ventilator usage pattern in the HiRID-II dataset, in terms of days on which a particular number of ventilators is used. **b.** Performance of RMS compared with the baseline, in terms of mean absolute error (MAE) for predicting the maximum number of used ventilators at a fixed horizon in the future. **c**. Example of a typical ICU setting shown for a duration of one day, annotated with ground-truth, RMS and baseline predictions. In the rug plot, relevant better predictions are marked for the offsets 1-4. **d.** RMS compared with the baseline, for different absolute differences of predictions (1-4) in the rows, for a prediction horizon of 8-16 hours in the future. The table entries denote the proportion of time points in which either RMS or the baseline is better by at least the difference of the row. **e.** Performance of RMS for different current ventilator ICU usage scenarios, ranging from low usage, to high usage, compared with the baseline, for a prediction horizon of 8-16 hours in the future. The number of time points falling into each bin is denoted in parentheses. **f.** Performance of RMS, by hour of the day when the prediction is performed, compared with the baseline, for a prediction horizon of 8-16 hours in the future.

We trained a meta-model using the four scores to predict the number of ventilators in use in the ICU at future time horizons every hour (4-8h, 4-12h, 8-12h, 8-16h, 16-24h; **Fig. 4b**). We compared it with a baseline that predicts that the number of non-elective patients requiring mechanical ventilation remains stable. We observed that the proposed model clearly outperforms this baseline in terms of mean absolute error (MAE), with the largest relative gain in longer prediction horizons (**Fig. 4b**). In 39% of time points the model’s predictions were at least two ventilators closer to ground-truth, for predicted ventilator use in 8-16 hours (**Fig. 4c/d**). RMS outperformed the baseline for the majority of ICU ventilator utilization scenarios (**Fig. 4e**) with the largest improvement over the baseline when the respirator use is below the maximum capacity (**Fig. 4e**) and for predictions of ventilator use during day hours (**Fig. 4f**).

### Explorative joint analysis of RMS scores throughout the ICU stay

We analyzed the relationship of the four RMS scores produced at each time point of the ICU stay by embedding the most important parameters for respiratory failure and extubation failure prediction (union of the top 10 variables identified for each task, current value feature) using t-distributed stochastic neighbor embedding (t-SNE^31^) with subsequent discretization into hexes. This approach produces a two-dimensional hex-map that defines subsets of comparable patient states that can be compared across different characteristics, i.e. between the panels for the hex. We observed that the space is divided into two distinct states, corresponding to time points when the patient is ventilated or not ventilated (**Fig. 5a**). The region of ventilated patients is further subdivided, with patients in the upper part being more likely to be ready-to-extubate (**Fig. 5b**). As expected, the ventilated and not ready-to-extubate region has the highest observed 24h mortality (**Fig. 5c**). Patients experiencing respiratory failure were concentrated in a compact region in the area corresponding to non-ventilated patients, as well as scattered throughout the area corresponding to ventilated patients (**Fig. 5d**). States with high risk of future ventilation need according to RMS-MV_Start_ are close to the boundary of the ventilated region (**Fig. 5e**). Readiness to extubate scores show a less clear pattern, but scores tend to be higher in the upper part of the ventilated region, which is also enriched in states in which patients are ready-to-extubate (**Fig. 5f**). For RMS-EF, high scores are concentrated in two distinct regions at the edge of the ventilated region (**Fig. 5g**). Lastly, RMS-RF scores are high close to the boundary of patients already in respiratory failure (**Fig. 5h**). The median risk scores of hexes for respiratory failure/ventilation need are strongly positively correlated with an R^2^ of 0.471 (**Fig. 5i**). Likewise, respiratory failure and extubation failure scores were moderately positively correlated (**Fig. 5j**). For RMS-EF/RMS-MV_End_ scores, no correlation could be observed (**Extended Data Fig. 10**). For three exemplary hexes with predominantly (1) non-ventilated patients but high RMS-RF score, (2) ready-to-extubate patients but high RMS-EF score, and (3) not-ready-to-extubate patients but high RMS-RF score, the distribution of clinical parameters was analyzed, showing plausible relationships with clinical parameters. For instance, the non-ventilated patients with the highest RF risks had low PaO_2_, high (supplemental) F_i_O_2_ and high respiratory rates (**Fig. 5k**).

**Fig. 5:**
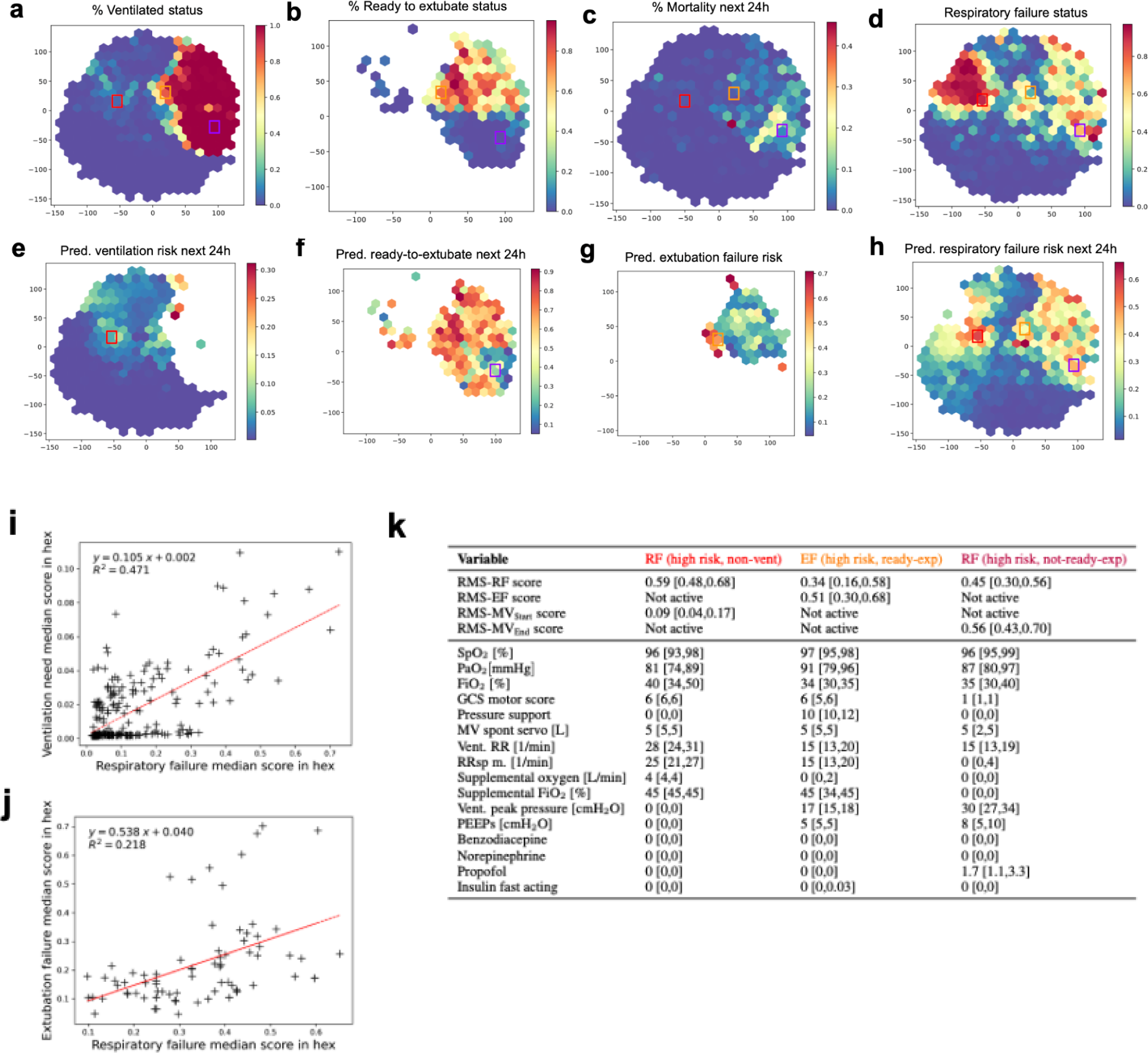
Joint analysis of the four scores produced by RMS overlaid on a t-SNE embedding based on important respiratory parameters. In all plots, x,y axes represent the two dimensions of the two-dimensional t-SNE embedding space. **a.** Hexes are colored by the proportion of time points in the hex for which the patient is ventilated. **b.** Hexes are colored by the proportion of time points in the hex for which the patient is ready-to-extubate given the patient is ventilated. **c.** Hexes are colored by observed 24h mortality risk. **d.** Hexes are colored by the proportion of time points in the hex for which the patient is in respiratory failure. **e-h.** The color of the hex denotes the median RMS-MV_Start_/RMS-MV_End_/RMS-EF/RMS-RF scores of the time points assigned to the hex, respectively. **i.** Relationship of median respiratory failure score (RMS-RF) and median ventilation need score (RMS-MV_Start_) in hexes for time points where both scores are active. The p-value of a Wald test for a non-zero regression line slope is 1.5*10^−36^. **j.** Relationship of median respiratory failure score (RMS-RF) and median extubation failure score (RMS-EF) in hexes for time points where both scores are active. The p-value of a Wald test for a non-zero regression line slope is 2.7*10^−5^. **k.** Score and input value distribution of time points assigned to three selected hexes for the 16 variables used as input for the t-SNE. The median is reported, and numbers in square brackets refer to the interquartile range.

## Discussion

We presented a ML-based system for the comprehensive monitoring of the respiratory state of ICU patients (RMS). RMS consists of four highly accurate scoring models that predict the occurrence of respiratory failure, start of mechanical ventilation, readiness to extubate as well as tracheal extubation failure. By combining the prediction scores of all admitted patients at any time point and by accounting for the likelihood of future admissions, RMS facilitated the accurate prediction of the near future cumulative number of patients requiring mechanical ventilation, which may help to optimize resource allocation within ICUs.

The HiRID-II dataset released alongside this work is a rich resource for broad-scale analyses of ICU patient data. It represents an important advance over HiRID-I, both in terms of the number of included patients and the number of clinical parameters that are included. Our initial analysis of the HiRID-II dataset identified clinically significant links; both the presence and duration of respiratory failure, as well as extubation failure, were associated with increased ICU mortality, indicating distinct yet interconnected risk factors. These insights highlight the critical need for advanced alarm systems for clinical settings to reduce the risks associated with respiratory and extubation failure. The release of the HiRID-II dataset on Physionet^18,19^ will offer numerous opportunities for further research, allowing for more in-depth investigations into various aspects of ICU patient care and outcomes.

RMS-RF predicts respiratory failure throughout the ICU stay, and alarms for impending failure were typically triggered at least 10 hours before the event. This early warning has the potential for clinicians to optimise medical therapy and potentially prevent the need for intubation. The transparent break-up of the model’s alarm output into SHAP values of the most relevant parameters may inform clinician understanding and guide their actions. RMS-RF outperformed a baseline representing standard clinical decision-making based on SpO_2_ and F_i_O_2_, and significantly reduced the number of false alarms. It produces RF-specific alarms and silences them within a specified period of time after the model triggers an alarm, reducing alarm fatigue, which is a major issue for ventilator alarms^32^. Prior to respiratory failure, only 1.5 alarms per patient/day were raised, which is manageable for the clinical personnel, and unlikely to cause alarm fatigue. Reassuringly, only variables directly associated with respiratory physiology or ventilator settings were found to be predictive of impending respiratory failure. RMS-RF demonstrated its highest precision in individuals admitted with cardiovascular or respiratory admission diagnoses, while its performance notably declined in neurologic patients. In these patients ventilatory management is often determined by the need to protect a compromised airway in patients with altered levels of consciousness and not by the presence of RF per se^33^. To date, few externally validated ML models continuously predicting acute respiratory failure in the ICU have been reported. Recent works by Le et al.^10^, Zeiberg et al.^34^, and Singhal et al.^35^ focus on mild respiratory failure (P/F index < 300 mmHg). Other models predict respiratory failure at the time of ICU admission or are only valid for specific cohorts^36–38^.

RMS-EF predicts tracheal extubation failure and significantly outperformed a clinical baseline derived from common clinical criteria for assessment of readiness to extubate status. The model was well calibrated, with almost ideal concordance of the prediction score and observed risk of extubation failure. A potential use case would be to assess the predicted failure risk to determine whether to accelerate or delay the extubation of the patient. At 80% recall, a quarter of correctly predicted extubation successes were recommended more than 3h before the actual extubation. This exceeds the recall of routinely used readiness tests and suggests that our model could help clinicians to extubate patients earlier. However, in our analysis we could not ascertain whether a patient was not extubated for reasons not apparent from the data, such as availability of staff. For clinical use the model could also be operated at 20% recall with very high precision (80%), to identify patients with a high likelihood that extubation will be unsuccessful. This could caution clinicians from prematurely extubating high-risk patients. For the prediction of extubation failure, various models have been proposed^39–44^. The largest cohorts to date were used in the works by Zhao et al.^44^, who only validated the model in a cardiac ICU cohort, which limits the generalizability of the results, and Chen et al.^45^, who restricted the evaluation to ROC-based metrics, and do not discuss the clinical implications of the model’s performance.

ML has previously been used to develop support systems for the management of RF patients in the ICU. These included models for detection of ARDS^7–11^ and COVID-19 pneumonitis patients^35,46^, prediction of readiness-to-extubate^47–49^, need for mechanical ventilation^50,51^, and detection of patient-ventilator asynchrony^52^. Existing work focused on single aspects of RF management, often in specific patient cohorts only. Our approach aims to comprehensively monitor the respiratory state throughout the RF treatment process, by integrating relevant respiratory-system related tasks and allowing for joint analysis of risk scores and trajectories. We believe a single and universally applicable system is much more likely to be successfully implemented than multiple fragmented models relating to specific disease entities. A further distinguishing feature of RMS is the five-minute time resolution at which predictions are made, enabling longitudinal analysis of risk trajectories. The dynamic prediction, which is a central feature of our model, is more flexible than traditional severity scores, which are evaluated at fixed time points, such as at 24 h after ICU admission^53^, mainly to predict ICU mortality^54^.

For successful external validation of RMS-RF, it was key to exclude medication variables from the model, as their inclusion was detrimental to model transferability. We hypothesize that this difficulty is caused by the observed medication policy differences between centers. Interestingly, ventilator settings, while also policy-dependent, did not compromise transfer performance in the same way. Investigating and quantifying the underlying policy differences, which made transfer difficult, needs additional research. Model transferability is an important topic in robust ML algorithms for ICU settings, where it has been recently studied in risk prediction in sepsis^15,55,56^ and mortality^57^. Our results suggest that medication variables require special attention to enable transfer. In contrast to RMS-RF, we suggest that RMS-EF to be re-trained and fine-tuned using the data from the center where it should be applied. The policy differences between different centers proved more detrimental to its performance than for RMS-RF.

Clinical prediction models for individual patients have been extensively studied. Resource planning in the ICU has received little attention in the ML literature, but came into renewed focus due to the COVID-19 crisis^58^. The first ML-based models to predict ICU occupancy were proposed during the pandemic. Lorenzen et al.^58^ predicted daily ventilator use as well as, more generally, hospitalization up to 15 days into the future^59^. The RMS presented here clearly outperformed a baseline method for predicting future ventilator use at the ICU level. With a mean absolute error of 0.39 ventilators per 10 ICU patient beds used during the next shift (8 to 16 hours), the model is sufficiently precise for practical purposes. Since resource allocation in the ICU depends on local policies and procedures, such a system likely needs to be retrained for every clinical facility for reliable predictions. External validation was not feasible as all public ICU datasets have random date offsets and therefore no information on concurrent patients in the ICU^60^.

In this study, we developed predictors of key aspects of respiratory state management, including RF, extubation failure, the need for mechanical ventilation, and readiness for tracheal extubation. These predictors collectively describe various aspects of a patient’s respiratory status in the ICU which can be used for exploratory analysis. The joint analysis and visualization of risk scores alongside other vital clinical variables yielded discernible clusters that correspond to specific patient states, indicating the potential for risk stratification within a patient population. We observed a separation of patient states into two main clusters that align with ventilated and non-ventilated states, with substructures within these clusters. The patients with highest 24h mortality risk identified on the hex-map often had depressed levels of consciousness, were more likely to require mandatory modes of mechanical ventilation, had higher peak airway pressure and required higher PEEP; all indicators of more severe underlying lung pathology. We also identified a cluster of patients who are clinically ready to be extubated, and have a low risk of RF but a very high extubation failure risk. These patients required relatively higher airway ventilation pressure and had a low respiratory rate, which are all established risk factors for extubation failure.

The hex-map visualization allows for the monitoring of individual patient states over time with updates, akin to those seen in methodologies like T-DPSOM^61,62^. This dynamic tracking is based on the automated integration of multiple respiratory state dimensions and uses nonlinear dimensionality reduction to provide the position of an individual patient on the map of respiratory health states. We expect that hex-map visualization has the potential to assist clinicians in identifying changes in patients’ clinical states, although the practical implications of this feature require further validation. This represents a different approach to previous work, that mainly tries to understand biological phenotypes of ARDS patients^63–66^ or longitudinal sub-phenotypes of a more specific patient set, like COVID-19 patients^67,68^. Overall, while the hex-map visualization provides an interesting perspective for monitoring of respiratory state in the ICU and can serve as a tool for a more detailed exploration, the presented analysis is exploratory only. Further research is needed to substantiate the clinical relevance of the identified clusters and to explore how this system might integrate into the decision-making processes within the ICU.

Our study tried to avoid certain limitations of retrospective model development studies. Unlike typical single-center studies, our research utilized data from two distinct centers, one for development and another for validation. This approach reduced the risk of overfitting models to a local patient cohort, although it is important to note that external applicability may still vary and retraining on local data will be needed for parts of the proposed RMS. We have incorporated improvements based on our previous work into our models. Unlike earlier systems that were heavily reliant on sporadic clinical measurements, such as serum lactate concentration^12^, our current model uses continuous SpO_2_ monitoring and ventilator data. Use of automated continuous data reduces the influence of clinician-driven decisions on our alarm systems, ensuring a more objective assessment of the patient’s condition. However, the retrospective nature of our data collection is still a limitation. Missing data was partially imputed for respiratory failure annotation, and while this aids in model development, it introduces potential biases. This study does not address how integrating the system into everyday clinical practice might influence treatment or monitoring strategies (a phenomenon known as domain shift^69^). Specifically, if the model would rely heavily on clinician-driven interventions (such as changes in PEEP or the administration of diuretics) as predictors of respiratory failure, any future alterations in clinician behavior (possibly driven by the model implementation) could reduce the model’s predictive accuracy. We constructed clinical baselines as best-effort reference points for comparison, derived from the data available in our cohort. As such, they are not established standards and may miss important clinical elements, such as respiratory effort, that are not routinely recorded but available to physicians in practice. Lastly, our assessment of the extubation failure risk score was limited to scenarios of actual extubation events. While we think that the accuracy of this score would be similar in patients nearing readiness for extubation, this cannot be definitively concluded from our retrospective data. Future prospective implementation studies are needed to fully understand the implications of our model in a live clinical setting.

In summary, we have developed a comprehensive monitoring system for the entire respiratory failure management process. We have shown that our system has the potential to facilitate early identification and assessment of deteriorating patients, aiming to enable rapid treatment; and to simplify resource planning within the ICU environment. The physiological relationship between risks and individual predictions can be inspected using SHAP values, thereby hopefully offering valuable insights to clinicians, and ultimately increasing trust in the system^70^. The potential benefit of the system in improving patient outcomes needs to be validated in prospective clinical implementation trials.

## Data Availability

Data used in this study were obtained from University Hospital Bern and we aim to make it available on physionet.org in anonymized form, similar to the HIRID-I dataset associated with a previous study (Hyland et al., Nature Medicine, 2020).

## Acknowledgments

This project was supported by the Grant No. 205321_176005 of the Swiss National Science Foundation (to T.M.M. and G.R.), and grant #2022-278 of the Strategic Focus Area “Personalized Health and Related Technologies (PHRT)” of the ETH Domain (Swiss Federal Institutes of Technology), and ETH core funding (to G.R.). We acknowledge discussions with and organizational, administrative or technical help by Carmen Pfortmüller, Jörg Schefold, Daniel Vonder Mühll, Olga Mineeva, Quinten Johnson, Dinara Veshchezerova, David Meyer, Anastasia Escher, Nora Toussaint, Margarita Kuznetsova, Fedor Sergeev, Marc Zimmermann, Catherine Jutzeler, Karsten Borgwardt, Thomas Gumbsch, Bowen Fan, Jörg Goldhahn, Sonia Strangio, Ivo Schauwecker, Martina Baumann, Sergio Maffioletti, Bernd Rinn, Anna Wiegand, Diana Coman Schmidt, Matthew Levin, Robert Freeman, Thomas Fuchs, Emanuela Keller, Michael Krauthammer, Paul Elbers, and Patrick Thoral. Computational analyses were performed at the LeonhardMed Trusted Research Environment at ETH Zurich (https://sis.id.ethz.ch/services/sensitiveresearchdata/). The work by S.L.H. was done while she was working at ETH Zurich. We thank David Sidebotham for proofreading the manuscript.

## Author contributions

M.H., X.L., M.F., G.R., T.M.M. with input from S.L.H., M.Ho., A.P., H.Y., M.B. designed the experiments; M.F., T.M.M., D.B. selected and provided the clinical data and context; A.P. with input from M.F., G.R., T.M.M., D.B., X.L. k-anonymized the dataset. X.L., M.F., A.P. with contributions from T.M.M., G.R., D.B. preprocessed and cleaned the HiRID-II data; X.L., M.F. harmonized the UMCdb dataset with HiRID-II. M.H., M.F. with input from G.R., T.M.M, X.L., S.L.H. defined and developed the respiratory state annotations and labels; M.H., M.F. developed the continuous estimation algorithm for PaO2;. M.H., M.F. developed and extracted ML features; M.H. developed the pipeline for supervised learning including variable selection; M.Ho. with input from X.L., G.R., M.F., M.H. performed the fairness analysis in sub-cohorts. A.P., with input from M.H., M.F., T.M.M., G.R., performed analyses of treatment policy differences. X.L. with input from G.R., M.H., M.F., A.P. conceived and developed the model for resource planning, M.H. with input from G.R., M.F., X.L., T.M.M. implemented the joint analysis of RMS scores; T.M.M., G.R., M.F. conceived and directed the project; M.H., M.F., X.L., G.R., T.M.M, A.P., M.Ho. with input from H.Y., M.B., D.B. wrote the manuscript. X.L. with input from all authors created Fig. 1. All authors read the manuscript and provided critical feedback.

## Extended data figures

**Extended Data Fig 1.**
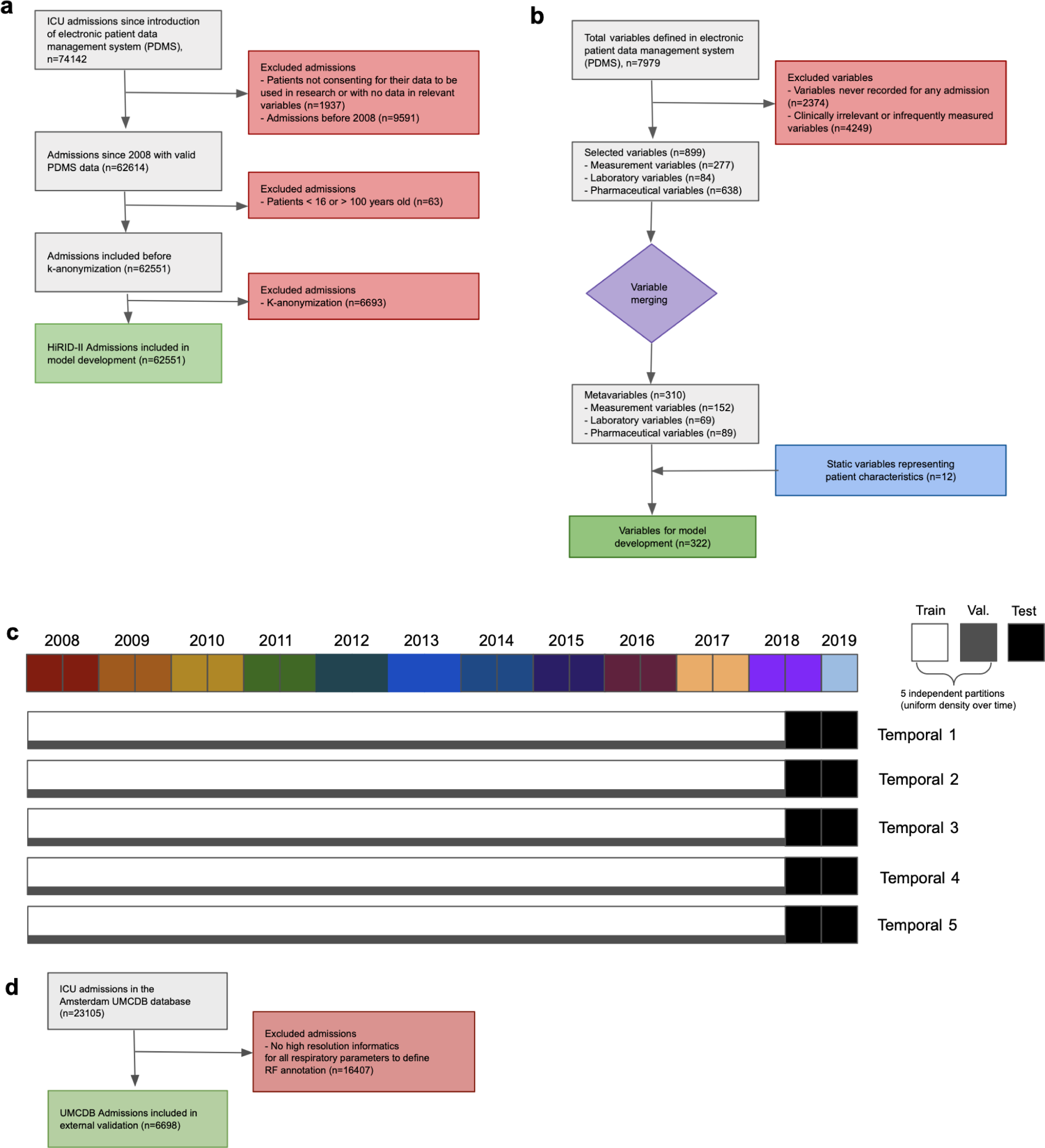
Patient inclusion & Experimental design. **a.** Patient inclusion schema in the HiRID-II dataset. **b.** Inclusion of clinical parameters in the data extraction pipeline of the HiRID-II dataset. **c.** Split design schema for performance evaluation. A fixed test set consisting of admissions starting in Mid June 2018 to the end of 2019 was used, which is shared by all five temporal splits, and is marked by a black block in all five splits. The remaining patients were randomly partitioned five times into a training and validation set, each defining a temporal split, which is indicated by the horizontal white and grey bars. **d.** Patient inclusion schema in the UMCdb dataset used for external validation.

**Extended Data Fig 2.**
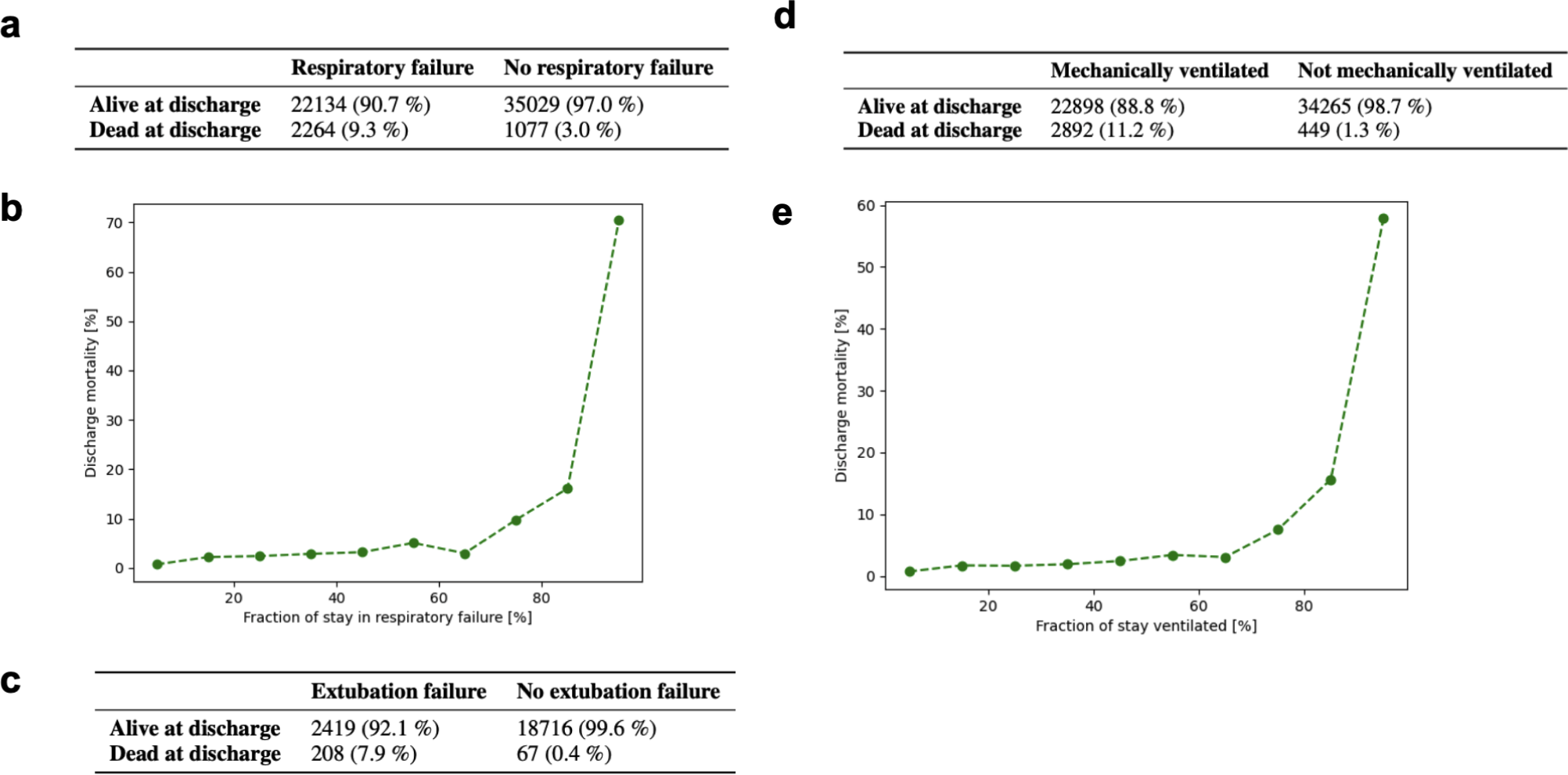
Association of ICU Mortality with Respiratory Failure / Extubation Failure / Ventilation. **a.** Mortality statistics for patients with respiratory failure at some time during their ICU stay, and those without respiratory failure during their ICU stay. **b.** Relationship of ICU mortality with fraction of the ICU stay in which patients experience respiratory failure. **c.** Mortality statistics for patients with extubation failure, and those without extubation failure but with at least one successful extubation. **d.** Mortality statistics for patients receiving mechanical ventilation during their ICU stay, and those not ventilated. **e.** Relationship of ICU mortality rate with fraction of their stay during which patients are mechanically ventilated.

**Extended Data Fig 3.**
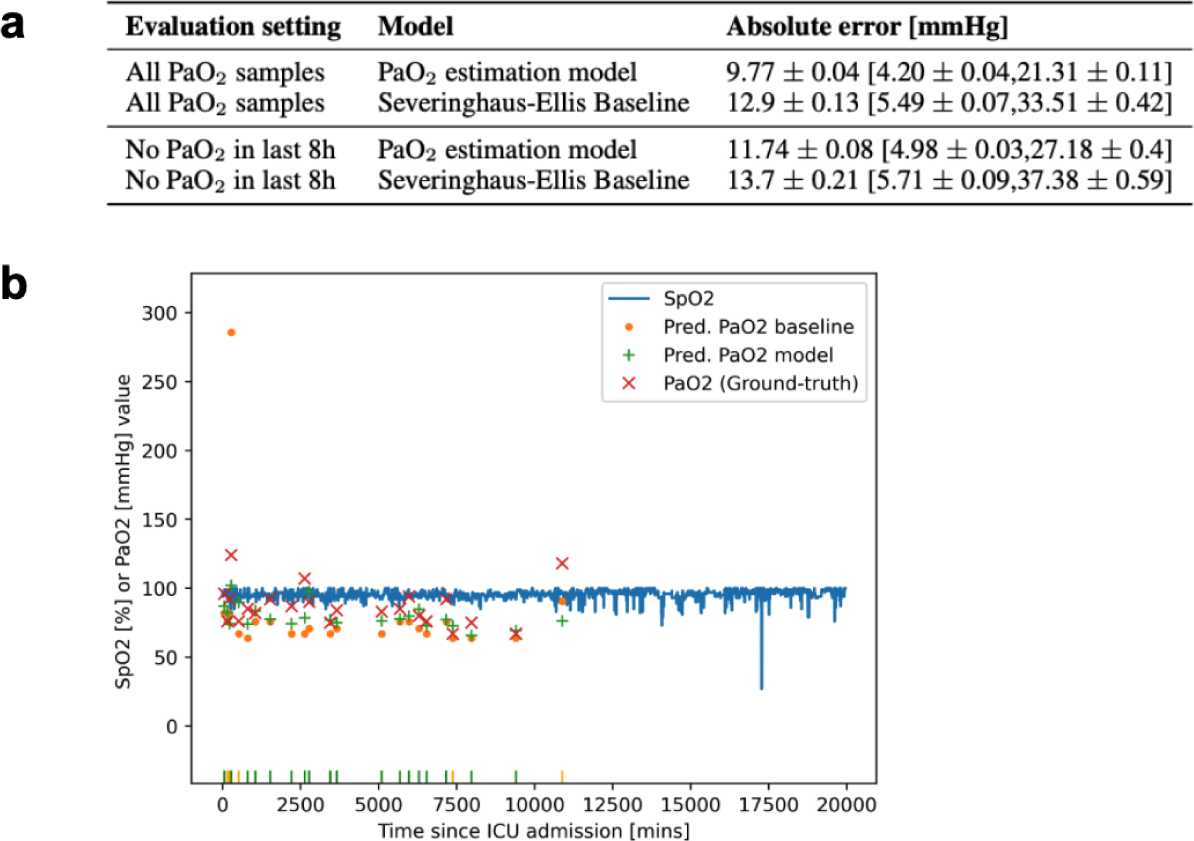
Performance of PaO_2_ estimation model. **a.** Performance evaluation of PaO_2_-estimation model on HiRID-II test set in terms of MAE vs. ground-truth PaO_2_ from invasive blood tests, compared with the non-linear Severinghaus-Ellis baseline. Error bars were obtained by re-sampling the test set with 50 %, 5 times at random, using complete patients. **b.** Example time series of predicted and ground-truth PaO_2_ values, as well as SpO_2_ values, and baseline predictions. A patient was selected at random for which the median absolute error of both model and baseline is close (within 1 mmHg) to their population median reported in panel a. The rug plot indicates time-points for which each model performs better than the other.

**Extended Data Fig 4.**
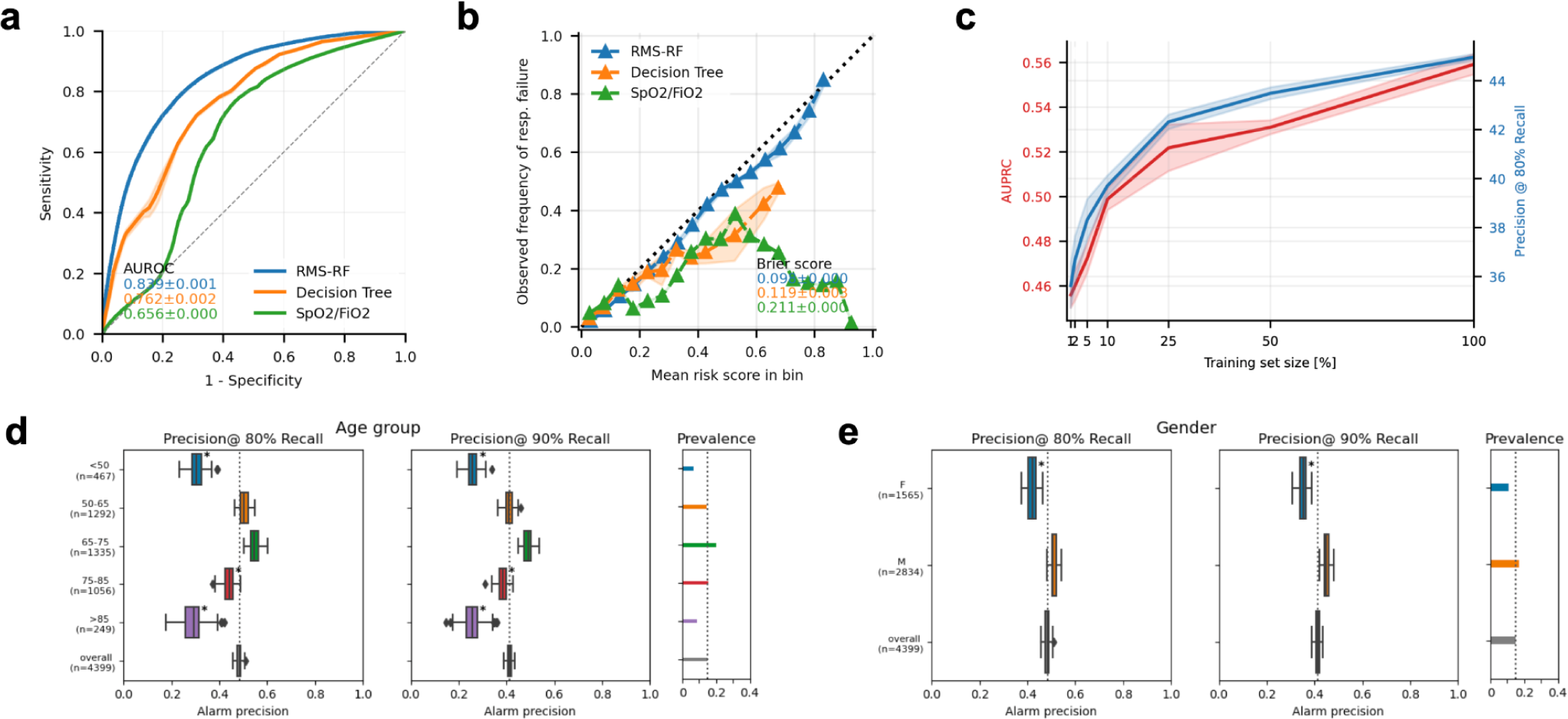
Evaluation of RMS-RF. **a.** ROC-based performance of the RMS-RF score, compared with the two baselines. **b.** Calibration of the RMS-RF model compared with the two baselines. **c.** Performance of the RMS-RF model, as the training set size is varied, in terms of complete patients **d.** Performance of the RMS-RF model by age group, for event recalls of 80/90 %. The model was re-calibrated for each sub-group using information available at admission time, to achieve a comparable event recall. When the prevalence of an event decreases, a greater proportion of positive results will be false, reducing the test’s precision. **e.** Performance of the RMS-RF model by gender, for event recalls of 80/90 %. The model was re-calibrated for each sub-group using information available at admission time, to achieve a comparable event recall.

**Extended Data Fig 5.**
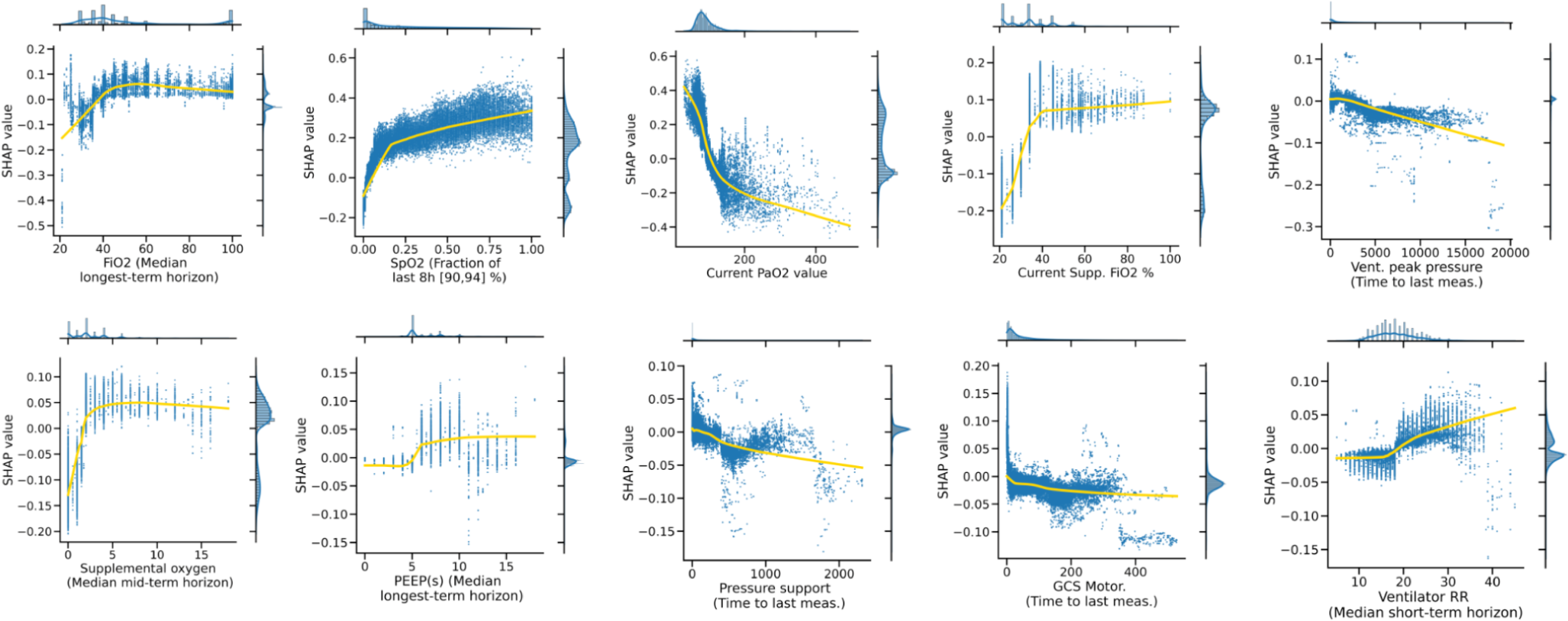
Model introspection of RMS-RF. SHAP value - feature value interactions of the top feature of the top 10 most important variables contained in RMS-RF.

**Extended Data Fig 6.**
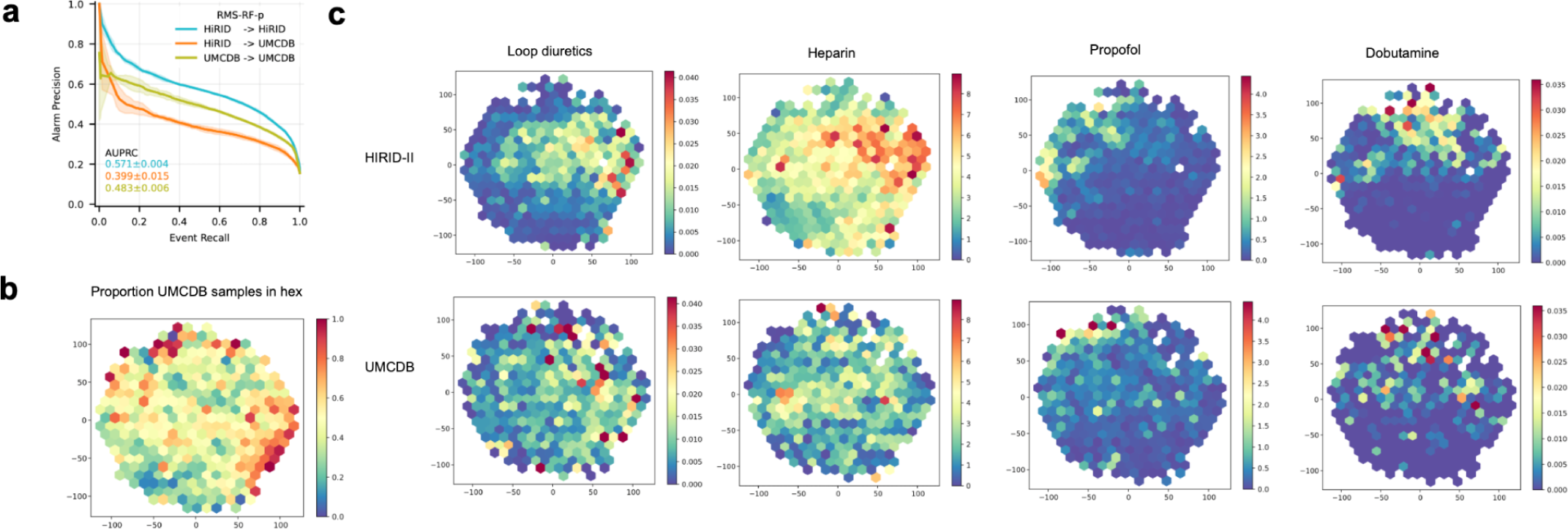
External validation of RMS-RF-p / Medication policy comparison HiRID-II/UMCdb. **a.** Performance of the RMS-RF-p model, which additionally includes medication variables, when trained and tested on HiRID-II, transferred to UMCdb and retrained in the UMCdb dataset **b.** t-SNE embedding of time points in the test set of a pooled dataset between samples from HiRID-II and UMCdb (1:1 ratio of two datasets), of physiological parameters. Only time points when the patient is not in respiratory failure are taken into account, for which the RMS-RF-p model is active. The color indicates the proportion of time points in the UMCdb dataset in a given hex. **c.** The same t-SNE embedding as in b is displayed separately for time points from the HiRID-II dataset, and the UMCdb dataset, corresponding to the rows. The hexes in the t-SNE are colored by the mean drug dosage of all time points assigned to the hex. The four medication variables, for which transfer issues of the RMS-RF-p model were detected, are analyzed in the columns. Medication policy differences are visible for all four variables, in particular for Heparin & Propofol.

**Extended Data Fig 7.**
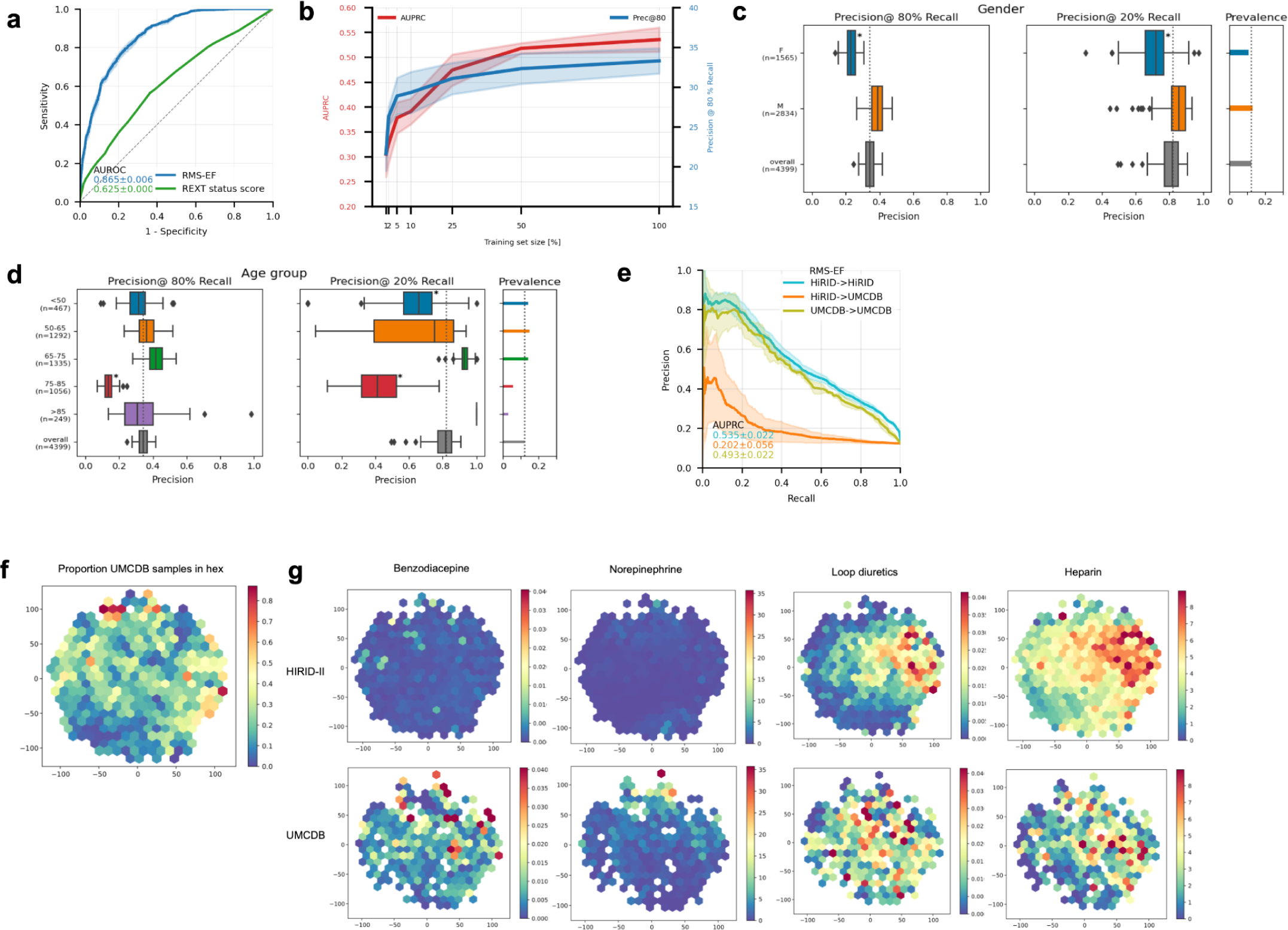
Evaluation of RMS-EF / Medication policy comparison HiRID-II/UMCdb. **a.** ROC-based performance of RMS-EF, compared with the baseline. **b.** Performance of RMS-EF as the training set size is varied between 1 % and 100 % of the original dataset size, by subsampling complete patient records in the training set. **c.** Performance of RMS-EF stratified by gender, at recall of 80/20 %. The model was re-calibrated for each sub-group using information available at the time of admission, to achieve a comparable recall. **d.** Performance of RMS-EF for different age groups, at recalls of 80/20 %. The model was re-calibrated for each sub-group using information available at the time of admission, to achieve a comparable recall. **e.** Performance of the RMS-EF model, when trained/tested in the HiRID-II dataset, transferred to the UMCdb dataset, and retrained in the UMCdb dataset. **f.** t-SNE embedding of time points in the test set of a pooled dataset between samples from HiRID-II and UMCdb (1:1 ratio of two datasets), of physiological input variables. Only time points when the patient is ready-to-extubate are taken into account, for which the RMS-EF model is active. The color indicates the proportion of time points in the UMCdb dataset in a given hex. **g.** The same t-SNE embedding as in g is displayed separately for time points from the HiRID-II dataset, and the UMCdb dataset, corresponding to the rows. The hexes in the t-SNE are colored by the mean drug dosage of all time points assigned to the hex. The four medication variables, for which transfer issues of the RMS-EF model were detected, are analyzed in the columns. Medication policy differences are visible for all four variables, in particular for Benzodiacepine & Norepinephrine.

**Extended Data Fig 8.**
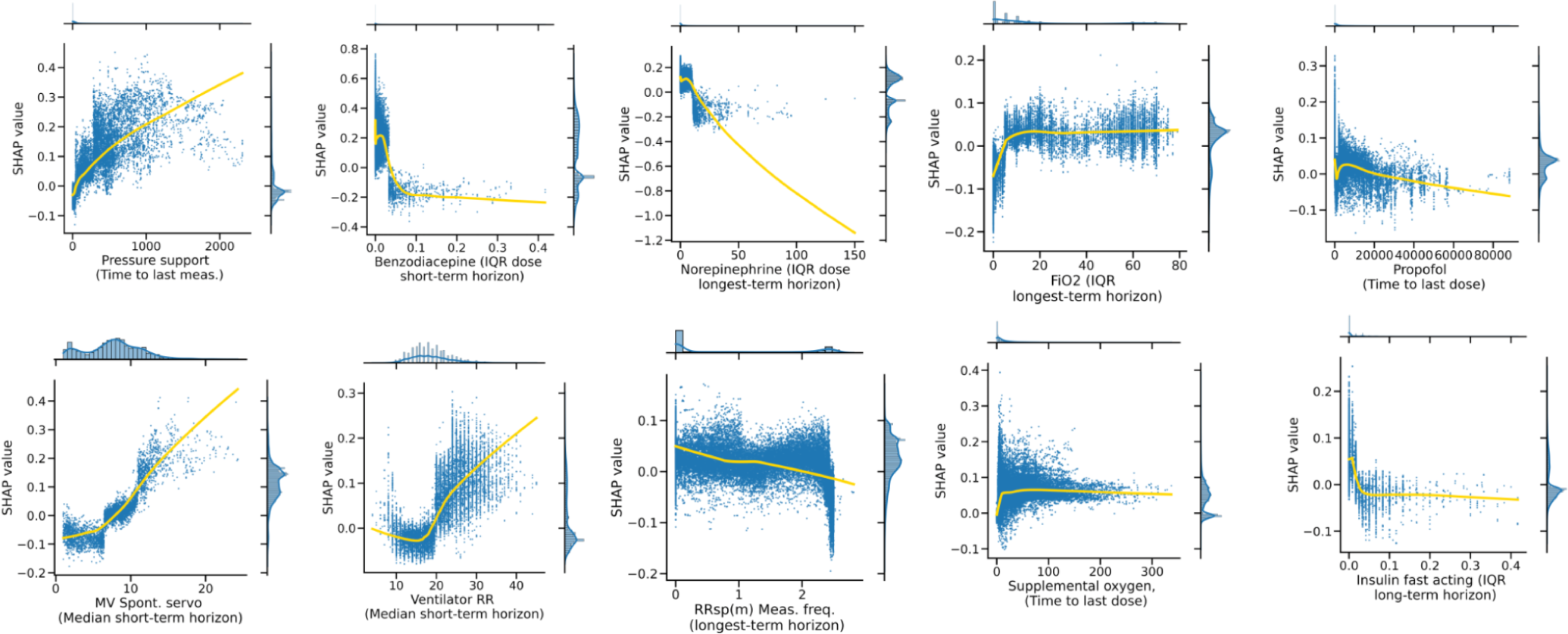
Model introspection of RMS-EF. SHAP value - feature value interactions for the top feature of the top 10 most important variables contained in the RMS-EF model.

**Extended Data Fig 9.**
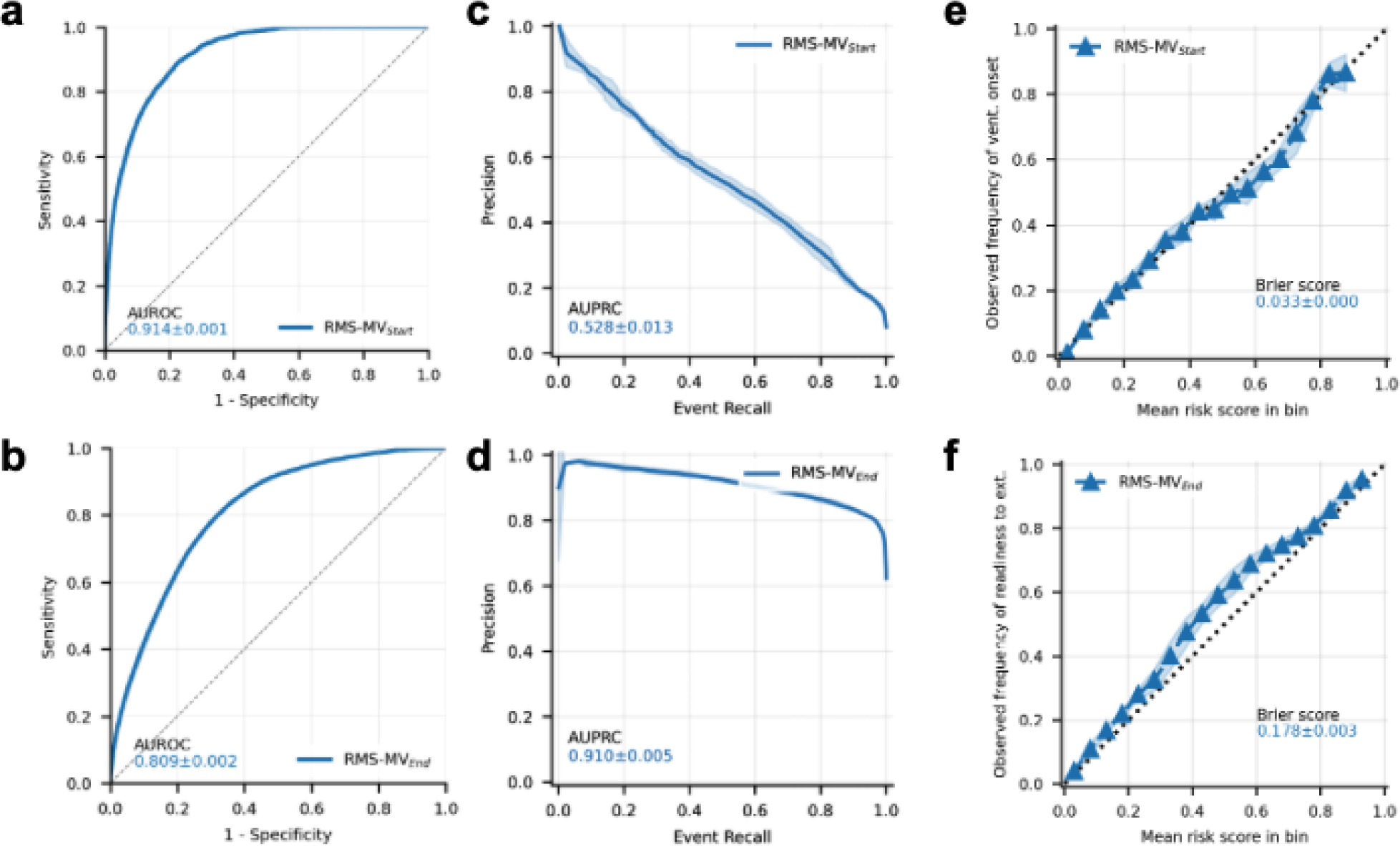
Evaluation of RMS-MV_Start_/RMS-MV_End_. **a.** ROC-based performance of RMS-MV_Start_, predicting ventilation onset within the next 24h. **b.** ROC-based performance of RMS-MV_end_, predicting being newly ready to extubate within the next 24h. **c.** Event-based PRC of the RMS-MV_Start_ alarm system. **d.** Event-based PRC of the RMS-MV_End_ alarm system. **e.** Calibration of the RMS-MV_Start_ score. **f.** Calibration of the RMS-MV_End_ score.

**Extended Data Fig 10.**
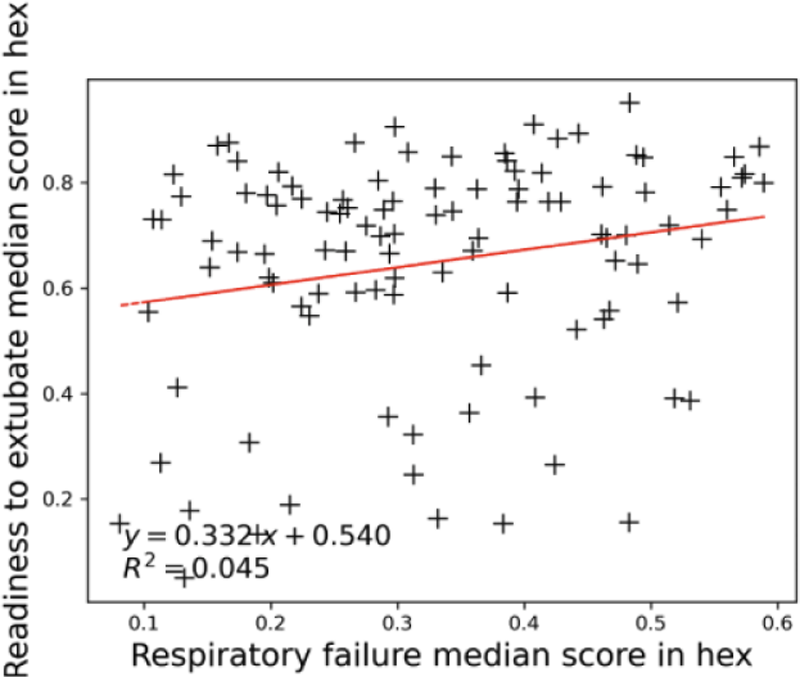
Joint task analysis details. Scatter plot of median respiratory failure vs. median readiness to extubate score in the hexes analyzed in the explorative joint analysis of RMS scores (see Fig. 5). A light positive correlation between respiratory failure and readiness to extubate scores can be observed, which is barely significant at 5 % level. A Wald test which tests non-zero slope of the regression line (shown in red) was performed.

## Supplemental Materials

**Supplemental Table 1.** Details on the clinical parameters extracted in the HiRID-II dataset (downloadable XLSX file).

**Supplemental Table 2.** Details on the imputation parameters, such as normal value, and imputation models, for the clinical parameters (downloadable XLSX file).

**Supplemental Table 3.** List of important variables used for computing complex features, as a basis for variable selection, and for building the final models RMS-RF/RMS-EF/RMS-MV_Start_/RMS-MV_End_ (downloadable XLSX file).

**Supplemental Table 4.** List of severity levels for computing ‘instability history’ features, for a subset of the important variables. (downloadable XLSX file).

**Supplemental Table 5.** Model training parameters and grid used for selection of hyperparameters for the LightGBM library (downloadable XLSX file).

## Footnotes

1 We currently work on the approvals for the release of the newer dataset and expect to have it ready at time of publication of the manuscript.

## References

1. Vincent, J. L., Sakr, Y. & Ranieri, V. M. Epidemiology and outcome of acute respiratory failure in intensive care unit patients. Crit. Care Med. 31, S296–9 (2003).

2. Bellani, G. et al. Epidemiology, Patterns of Care, and Mortality for Patients With Acute Respiratory Distress Syndrome in Intensive Care Units in 50 Countries. JAMA 315, 788–800 (2016).

3. Vincent, J.-L. et al. The epidemiology of acute respiratory failure in critically ill patients(*). Chest 121, 1602–1609 (2002).

4. Donchin, Y. & Seagull, F. J. The hostile environment of the intensive care unit. Curr. Opin. Crit. Care 8, 316–320 (2002).

5. Sanchez-Pinto, L. N., Luo, Y. & Churpek, M. M. Big Data and Data Science in Critical Care. Chest 154, 1239–1248 (2018).

6. Wung, S.-F., Malone, D. C. & Szalacha, L. Sensory Overload and Technology in Critical Care. Crit. Care Nurs. Clin. North Am. 30, 179–190 (2018).

7. Bai, Y., Xia, J., Huang, X., Chen, S. & Zhan, Q. Using machine learning for the early prediction of sepsis-associated ARDS in the ICU and identification of clinical phenotypes with differential responses to treatment. Front. Physiol. 13, 1050849 (2022).

8. Lam, C., Thapa, R., Maharjan, J. & Rahmani, K. Multitask Learning With Recurrent Neural Networks for Acute Respiratory Distress Syndrome Prediction Using Only Electronic Health Record Data: Model …. JMIR Medical (2022).

9. Wu, J. et al. Early prediction of moderate-to-severe condition of inhalation-induced acute respiratory distress syndrome via interpretable machine learning. BMC Pulm. Med. 22, 193 (2022).

10. Le, S. et al. Supervised machine learning for the early prediction of acute respiratory distress syndrome (ARDS). J. Crit. Care 60, 96–102 (2020).

11. Ding, X.-F. et al. Predictive model for acute respiratory distress syndrome events in ICU patients in China using machine learning algorithms: a secondary analysis of a cohort study. J. Transl. Med. 17, 326 (2019).

12. Hyland, S. L. et al. Early prediction of circulatory failure in the intensive care unit using machine learning. Nat. Med. 26, 364–373 (2020).

13. Desautels, T. et al. Prediction of Sepsis in the Intensive Care Unit With Minimal Electronic Health Record Data: A Machine Learning Approach. JMIR Med Inform 4, e28 (2016).

14. Wang, D. et al. A Machine Learning Model for Accurate Prediction of Sepsis in ICU Patients. Front Public Health 9, 754348 (2021).

15. Moor, M. et al. Predicting sepsis using deep learning across international sites: a retrospective development and validation study. EClinicalMedicine 62, 102124 (2023).

16. Tomašev, N. et al. A clinically applicable approach to continuous prediction of future acute kidney injury. Nature 572, 116–119 (2019).

17. Faltys, M. et al. HiRID, a high time-resolution ICU dataset. PhysioNet 10.13026/NKWC-JS72 (2021).

18. Goldberger, A. L. et al. PhysioBank, PhysioToolkit, and PhysioNet: components of a new research resource for complex physiologic signals. Circulation 101, (2000).

19. Moody, G. B., Mark, R. G. & Goldberger, A. L. PhysioNet: a web-based resource for the study of physiologic signals. IEEE Eng. Med. Biol. Mag. 20, 70–75 (2001).

20. Sweeney, L. K-anonymity: A model for protecting privacy. Int. J. Uncertainty Fuzziness Knowledge Based Syst. 10, 557–570 (2002).

21. Thoral, P. J. et al. Sharing ICU Patient Data Responsibly Under the Society of Critical Care Medicine/European Society of Intensive Care Medicine Joint Data Science Collaboration: The Amsterdam University Medical Centers Database (AmsterdamUMCdb) Example. Crit. Care Med. 49, e563–e577 (2021).

22. Ke, G. et al. LightGBM: A Highly Efficient Gradient Boosting Decision Tree. in Advances in Neural Information Processing Systems 30 3146–3154 (2017).

23. Yèche, H. et al. HiRID-ICU-Benchmark --- A Comprehensive Machine Learning Benchmark on High-resolution ICU Data. Proceedings of the Neural Information Processing Systems Track on Datasets and Benchmarks 1, (2021).

24. Patel, S., Jose, A. & Mohiuddin, S. S. Physiology, Oxygen Transport And Carbon Dioxide Dissociation Curve. in StatPearls (StatPearls Publishing, Treasure Island (FL), 2023).

25. Wagner, P. D. Blood Gas Transport: Carriage of Oxygen and Carbon Dioxide in Blood. Semin. Respir. Crit. Care Med. 44, 569–583 (2023).

26. Pandharipande, P. P. et al. Derivation and validation of Spo2/Fio2 ratio to impute for Pao2/Fio2 ratio in the respiratory component of the Sequential Organ Failure Assessment score. Crit. Care Med. 37, (2009).

27. Brown, S. M. et al. Nonlinear Imputation of Pao2/Fio2 From Spo2/Fio2 Among Patients With Acute Respiratory Distress Syndrome. Chest 150, 307–313 (2016).

28. Hoche, M., Mineeva, O., Burger, M., Blasimme, A. & Rätsch, G. FAMEWS: a Fairness Auditing tool for Medical Early-Warning Systems. (2024).

29. Lundberg, S. M. & Lee, S.-I. A Unified Approach to Interpreting Model Predictions. in Advances in Neural Information Processing Systems 30 (eds. Guyon, I. et al.) 4765–4774 (Curran Associates, Inc., 2017).

30. Lundberg, S. M., Erion, G. G. & Lee, S.-I. Consistent Individualized Feature Attribution for Tree Ensembles. arXiv [cs.LG] (2018).

31. Maaten, L. van der & Hinton, G. Visualizing data using t-SNE. J. Mach. Learn. Res. 9, 2579–2605 (2008).

32. Scott, J. B., De Vaux, L., Dills, C. & Strickland, S. L. Mechanical Ventilation Alarms and Alarm Fatigue. Respir. Care 64, 1308–1313 (2019).

33. Cinotti, R. et al. Extubation in neurocritical care patients: the ENIO international prospective study. Intensive Care Med. 48, 1539–1550 (2022).

34. Zeiberg, D. et al. Machine learning for patient risk stratification for acute respiratory distress syndrome. PLoS One 14, e0214465 (2019).

35. Singhal, L. et al. eARDS: A multi-center validation of an interpretable machine learning algorithm of early onset Acute Respiratory Distress Syndrome (ARDS) among critically ill adults with COVID-19. PLoS One 16, e0257056 (2021).

36. Bolourani, S. et al. A Machine Learning Prediction Model of Respiratory Failure Within 48 Hours of Patient Admission for COVID-19: Model Development and Validation. J. Med. Internet Res. 23, e24246 (2021).

37. Ferrari, D. et al. Machine learning in predicting respiratory failure in patients with COVID-19 pneumonia-Challenges, strengths, and opportunities in a global health emergency. PLoS One 15, e0239172 (2020).

38. Bendavid, I. et al. A novel machine learning model to predict respiratory failure and invasive mechanical ventilation in critically ill patients suffering from COVID-19. Sci. Rep. 12, 10573 (2022).

39. Igarashi, Y. et al. Machine learning for predicting successful extubation in patients receiving mechanical ventilation. Front. Med. 9, 961252 (2022).

40. Huang, K.-Y., et al. A machine learning model for prediction of successful extubation in patients admitted to the intensive care unit. (2022).

41. Zeng, Z., Tang, X., Liu, Y., He, Z. & Gong, X. Interpretable recurrent neural network models for dynamic prediction of the extubation failure risk in patients with invasive mechanical ventilation in the intensive care unit. BioData Min. 15, 21 (2022).

42. Wang, H. et al. Early prediction of noninvasive ventilation failure after extubation: development and validation of a machine-learning model. BMC Pulm. Med. 22, 304 (2022).

43. Otaguro, T. et al. Machine Learning for Prediction of Successful Extubation of Mechanical Ventilated Patients in an Intensive Care Unit: A Retrospective Observational Study. J. Nippon Med. Sch. 88, 408–417 (2021).

44. Zhao, Q.-Y. et al. Development and Validation of a Machine-Learning Model for Prediction of Extubation Failure in Intensive Care Units. Front. Med. 8, 676343 (2021).

45. Chen, T. et al. Prediction of Extubation Failure for Intensive Care Unit Patients Using Light Gradient Boosting Machine. IEEE Access 7, 150960–150968 (2019).

46. Lazzarini, N., Filippoupolitis, A., Manzione, P. & Eleftherohorinou, H. A machine learning model on Real World Data for predicting progression to Acute Respiratory Distress Syndrome (ARDS) among COVID-19 patients. PLoS One 17, e0271227 (2022).

47. Sayed, M., Riaño, D. & Villar, J. Predicting Duration of Mechanical Ventilation in Acute Respiratory Distress Syndrome Using Supervised Machine Learning. J. Clin. Med. Res. 10, (2021).

48. Jia, Y., Kaul, C., Lawton, T., Murray-Smith, R. & Habli, I. Prediction of weaning from mechanical ventilation using Convolutional Neural Networks. Artif. Intell. Med. 117, 102087 (2021).

49. Zeng, L. et al. VentSR: A Self-Rectifying Deep Learning Method for Extubation Readiness Prediction. in 2022 IEEE International Conference on Bioinformatics and Biomedicine (BIBM) 1369–1374 (2022).

50. Shashikumar, S. P. et al. Development and Prospective Validation of a Deep Learning Algorithm for Predicting Need for Mechanical Ventilation. Chest 159, 2264–2273 (2021).

51. Siu, B. M. K., Kwak, G. H., Ling, L. & Hui, P. Predicting the need for intubation in the first 24 h after critical care admission using machine learning approaches. Sci. Rep. 10, 20931 (2020).

52. Sottile, P. D., Albers, D., Higgins, C., Mckeehan, J. & Moss, M. M. The Association Between Ventilator Dyssynchrony, Delivered Tidal Volume, and Sedation Using a Novel Automated Ventilator Dyssynchrony Detection Algorithm. Crit. Care Med. 46, e151–e157 (2018).

53. Lapp, L., Roper, M., Kavanagh, K., Bouamrane, M.-M. & Schraag, S. Dynamic Prediction of Patient Outcomes in the Intensive Care Unit: A Scoping Review of the State-of-the-Art. J. Intensive Care Med. 38, 575–591 (2023).

54. Calvert, J. et al. Using electronic health record collected clinical variables to predict medical intensive care unit mortality. Ann Med Surg (Lond) 11, 52–57 (2016).

55. Roggeveen, L. et al. Transatlantic transferability of a new reinforcement learning model for optimizing haemodynamic treatment for critically ill patients with sepsis. Artif. Intell. Med. 112, 102003 (2021).

56. Chen, Q. et al. Transferability and interpretability of the sepsis prediction models in the intensive care unit. BMC Med. Inform. Decis. Mak. 22, 343 (2022).

57. Alves, T., Laender, A., Veloso, A. & Ziviani, N. Dynamic Prediction of ICU Mortality Risk Using Domain Adaptation. in 2018 IEEE International Conference on Big Data (Big Data) 1328–1336 (ieeexplore.ieee.org, 2018).

58. Lorenzen, S. S. et al. Using machine learning for predicting intensive care unit resource use during the COVID-19 pandemic in Denmark. Sci. Rep. 11, 18959 (2021).

59. Tariq, A. et al. Patient-specific COVID-19 resource utilization prediction using fusion AI model. NPJ Digit Med 4, 94 (2021).

60. de Kok, J. W. T. M. et al. A guide to sharing open healthcare data under the General Data Protection Regulation. Sci. Data 10, 1–10 (2023).

61. Manduchi, L., Hüser, M., Vogt, J., Rätsch, G. & Fortuin, V. DPSOM: Deep Probabilistic Clustering with Self-Organizing Maps. (2019).

62. Fortuin, V., Hüser, M., Locatello, F., Strathmann, H. & Rätsch, G. SOM-VAE: Interpretable Discrete Representation Learning on Time Series. (2018).

63. Calfee, C. S. et al. Subphenotypes in acute respiratory distress syndrome: latent class analysis of data from two randomised controlled trials. Lancet Respir Med 2, 611–620 (2014).

64. Alipanah, N. & Calfee, C. S. Phenotyping in acute respiratory distress syndrome: state of the art and clinical implications. Curr. Opin. Crit. Care 28, 1–8 (2022).

65. Wilson, J. G. & Calfee, C. S. ARDS Subphenotypes: Understanding a Heterogeneous Syndrome. Crit. Care 24, 102 (2020).

66. Bos, L. D. et al. Identification and validation of distinct biological phenotypes in patients with acute respiratory distress syndrome by cluster analysis. Thorax 72, 876–883 (2017).

67. Bos, L. D. J. et al. Longitudinal respiratory subphenotypes in patients with COVID-19-related acute respiratory distress syndrome: results from three observational cohorts. Lancet Respir Med 9, 1377–1386 (2021).

68. Joshi, R. & Szolovits, P. Prognostic physiology: modeling patient severity in Intensive Care Units using radial domain folding. AMIA Annu. Symp. Proc. 2012, 1276–1283 (2012).

69. Liley, J., et al. Model updating after interventions paradoxically introduces bias. (2020).

70. Stenwig, E., Salvi, G., Rossi, P. S. & Skjærvold, N. K. Comparative analysis of explainable machine learning prediction models for hospital mortality. BMC Med. Res. Methodol. 22, 53 (2022).

