## Supplementary methods for "RMS: A ML-based system for ICU Respiratory Monitoring and Resource Planning"

### Study design and setting

The study was designed as a retrospective cohort study to develop and validate a set of clinical prediction models that are combined to form a ML-based respiratory monitoring system. The study was performed using data from the Department of Intensive Care Medicine at the University Hospital Bern, an interdisciplinary unit admitting > 6,500 patients per year and the sole provider of intensive care for adults at this hospital. This dataset (HiRID-II) was used for model development and internal validation. For external validation, an open-source dataset from the Amsterdam University Medical Center, referred to as UMCdb[1], was used, and harmonized to match the same structure of the HiRID-II dataset.

##### Ethical approval and patient consent

The competent ethics committee (CEC) of the Canton of Bern approved the study (BASEC 2016 01463). The need for obtaining informed patient consent for patient data from the University Hospital of Bern was waived due to the retrospective and observational nature of the study and the use of anonymized data only. No IRB or CEC approval is required for the anonymous public external validation dataset from Amsterdam University Medical Center (BASEC Req-2024-00250).

##### Participants and data sources

Details about participants and patient inclusion criteria in the two datasets are described in Table 1 / Extended Data Figure 1.

### HiRID-II data

For this work, we prepared the second version of the High time Resolution Intensive Care Unit Dataset (HiRID-II), consisting of high-temporal-resolution data from over 55,000 patient admissions to the intensive care units (ICUs) at the Bern University Hospital in Switzerland between January 2008 and June 2019.

HiRID-II is an improvement and update of the first HiRID dataset released by Faltys et al. on Physionet[2], which contains over 33,000 patient admissions dating between January 2008 and August 2016. HIRID-II additionally includes patients without data for determining circulatory failure or receiving any form of full mechanical circulatory support (previously excluded from HiRID-I) and patient data between August 2016 and June 2019. The final dataset was obtained after applying exclusion criteria to 74,142 initial admissions (see flow chart in Extended Data Figure 1).

HiRID-I includes 681 variables recorded in the patient data management system (PDMS, GE Centricity Critical Care, General Electrics) which were merged into 209 meta-variables based on their clinical concepts. HIRID-II records 218 variables more than the HiRID-I dataset, and contains 113 more meta-variables after variable merging. It is planned to release a version of HiRID-II on Physionet that includes the new admissions and variables. Details about meta-variables are listed in Supplemental Table 1.

#### Anonymization procedure

To ensure the anonymization of individuals in the dataset, we followed the same procedures that were applied to the MIMIC-III and AmsterdamUMCdb datasets; which adopted the Health Insurance Portability and Accountability Act (HIPAA) Safe Harbor requirements and, in the case of AmsterdamUMCdb, also the European Union's General Data Protection Regulation (GDPR) standards[3,4].

This included the removal of all eighteen identifying data elements listed in HIPAA. Free text was removed from the dataset. Patient age, height and weight were grouped into bins of size 5, with patients aged 90 years and older binned together. K-anonymization was subsequently applied to the patient's age, weight, height and sex. This procedure was separately applied to the original HIRID-I dataset (anonymized by Faltys et al.), including the additional set of training patients (2008-2018), and the held-out test set (2018-2019).

Within these temporally distinct training and test sets, admission dates were shifted by a random offset to lie between 2100 and 2200, while preserving seasonality, time of day and day of week. Measurements and medications with changing units over time were standardized to the latest unit used, to ensure that the admission time point could not be deduced from the units used.

#### Data splits

The publicly-released temporal data split into development and test set was used as a basis for designing the data splits in HiRID-II; after implementing the K-anonymization procedure described above. The test set of this split was held-out and not used prior to generating the final results to avoid subtle overfitting during the model design process. The development set was further divided randomly by complete patients, allocating 80 % of the patients to the final training set, and 20 % of the patients to the validation set. The final training set was used for model training. The validation set was used for selecting optimal hyperparameters, as well as early stopping of the training process. Performance in the validation set guided model design decisions in the prototyping phase as well as selection of clinical parameters using greedy forward selection. This splitting procedure into training and validation sets was repeated independently 5 times, to produce 5 splits.

The model development dataset drawn from HiRID-II contains 51,457 patients and the test set 4,401 patients. A temporal splitting strategy analogous to the one presented by Hyland et al.[5] was used, with one fixed test set to minimize leakage of admission time information. Five independent partitions into the training and validation sets were extracted from the development dataset, to estimate variation of model performance, containing 41,165 and 10,292 patients respectively.

For UMCdb a fixed test set consisting of a 25 % random sample of patients was drawn. The remaining patients formed the development dataset, which was partitioned by five random splits in proportion 80:20 % into training and validation sets. All five training/validation splits shared the same above-mentioned test set.

#### Analysis platform

After extraction of the HiRID-II and UMCdb data all computational analyses were performed on a secure compute cluster environment located at ETH Zürich (https://scicomp.ethz.ch/wiki/Leonhard). Python3, with numpy, pandas, matplotlib and scikit-learn, formed the backbone of the data-processing pipeline. The LightGBM[6] package was used for model training. Processing was performed in a batched form across most steps of the processing pipeline, with the HiRID-II data set being split into 100 batches, and the UMCdb data set being split into 50 batches.

#### Data preprocessing, variable merging, artifact rejection, medication preprocessing

Similar to the preprocessing steps presented by Hyland et al.[5], we first removed different types of artifacts in the data, such as timestamp artifacts and variable-misnaming artifacts, out-of-range-value artifacts, and record duplication artifacts. For variables that encode cumulative values, we converted the cumulative values to a rate. The dose values of medication variables were either converted to a rate or a binary indicator depending on their clinical relevance to respiratory failure defined by the clinicians. Drugs that are given in the form of discrete boluses were converted to continuous rates over a defined duration of action. The duration of action differs for different drugs and the details can be found in Supplemental Table 1.

After artifact removal and converting relevant variables to rates, we merged variables with the identical or near-identilcal clinical meaning/function into one single meta-variable (i.e. core body temperature, rectal temperature, and axillary temperature, were merged to form the meta-variable temperature). Similarly, drugs with identical or equivalent compounds were merged into one medication group. For each meta-variable, we took the median value of the available measurements at each timepoint when any of the corresponding variables is measured.

#### Data imputation

A time grid with step size of 5-minute was used; with the admission time period defined as the time between the first and last heart rate measurement of the intensive care unit stay. The analyzed admission time was limited to the first 28 days of an individual ICU stay. ICU stays of more than 28 days are rare and using such data would potentially introduce a bias in the model development process.

At each grid point a binary measurement indicator column was introduced as 1 if there was an observation in the corresponding 5 minutes, and 0 otherwise. Further, a time to last measurement column was introduced as -1 if there is no previous observation prior to the grid point, or equal to the number of minutes since the last observation.

“Dense imputation” where every value on the grid is imputed with a finite value was used as pre-processing for endpoint annotation and label definition. In contrast, prior to feature extraction, data was gridded but only partially imputed, to preserve some missingness patterns in the data. Here a missing indicator (NAN) was left at grid points where the value could not be estimated in a clinically plausible way. Subsequently, we refer to the latter mode as “feature imputation”.

For each meta-variable in the HiRID-II data schema, an imputation algorithm was defined and applied. If there was no prior measurement before a grid point, the grid point was filled with a missing-value indicator (for “feature imputation”), or a clinically defined normal value (for “dense imputation”). If there was more than one observation with the same time-stamp, the mean of all such observations was used. The imputation modes were:

a) Indefinite forward filling: The last measurement was indefinitely forward filled to each later time point on the 5-minute grid.
b) Limited forward filling based on medical concepts: Each measurement was forward filled up to a maximum time of k minutes, with k manually specified based on clinical concepts.
c) Limited data-adaptive forward filling: The median and standard deviation of the observation intervals for a clinical parameter were estimated in the training set. Using these estimates, forward filling was applied for up to 2*median(interval) + IQR(interval). This method was used if the forward filling horizon cannot be specified based on medical concepts.
d) Attribute to exact grid point: No forward filling was used if a measurement is only relevant for a very short time at the exact grid point where it was observed. In this case, forward filling was limited to 5 minutes, i.e. only the grid point next to the measurement location contains the value.

The imputation modes used for each variable are listed in Supplemental Table 2, per clinical parameter.

For the variables ‘Cardiac output’, ‘Urine output’, ‘Fluid input’, ‘Fluid output’, imputation used special formulae to estimate the current value based on the patient’s observed/estimated height/weight and BMI values. More details are given in Supplementary Table 2.

For static variables, analogously to time series variables, ‘dense imputation’ guaranteed there are no missing values, and median/mode imputation based on statistics from the training set was used for continuous variables and categorical variables, respectively. ‘Feature imputation’ for static variables used no imputation, missing values were left as NAN.

#### $PaO_2$ estimation

The annotation of respiratory failure depends on the availability of a current $PaO_2$ value. To measure $PaO_2$, an arterial blood gas sample (ABGA) of the patient has to be drawn and processed. Therefore, $PaO_2$ measurements are only available at intervals determined by ABGA measurement frequency. For a continuous assessment of a patient’s respiratory state using P/F ratio, estimates of $PaO_2$ values have to be used when measurements are not available. In the clinical setting, continuously monitored pulse oximetry derived haemoglobin oxygen saturation ($SpO_2$) can be used to estimate the current $PaO_2$ value[7–9]. To reduce the effect of outliers, the $SpO_2$ time series was pre-processed with a percentile smoother (75% percentile kernel function, 30 min centralized kernel window). A literature review of existing models revealed that the non-linear parametric model by Ellis[8,9] performs best. We were able to further improve upon Ellis in $PaO_2$ estimation using 2 nested regularized L2 regression models by using 7 hand-crafted features, defined at each time-point of the stay. Polynomial features of degree 3 were then computed on these features to capture non-linear interactions explicitly. The following features were used:

- Last available $SpO_2$ measurement
- Last available $PaO_2$ measurement
- Last available $SaO_2$ measurement
- Last available pH measurement
- Time to last available $SpO_2$ measurement
- Time to last available $PaO_2$ measurement
- Closest $SpO_2$ measurement to the last $PaO_2$ measurement

This base model was nested into a meta-model which performed the final prediction. As input features the meta-model used (1) the same polynomial features of the base model, (2) the prediction made by the base model as well as (3) the prior mistakes, i.e. signed offsets between base model prediction and ground-truth of the (up to) 10 prior $PaO_2$ real measurements. This allows the model to adapt using the context of previous wrong predictions to improve its predictions over the time of the ICU admission, and adapt to the patient physiology at hand. Hereby the prediction of $PaO_2$ is independent of the $F_IO_2$, since complex physiological interactions between $F_IO_2$ and the P/F ratio exist, which may lead to unwanted mathematical coupling between our $PaO_2$ estimate and the P/F index.

The model was only trained using time-points where at least one prior measurement was available for each of $SpO_2$, $PaO_2$, $SaO_2$ and pH and a $PaO_2$ measurement was recorded at this time-point. The regression label was equal to the ground-truth $PaO_2$ measurement. During evaluation a prediction was only made if at least one $PaO_2$ measurement was previously observed. Before the first $PaO_2$ measurement, the normal imputation algorithm for $PaO_2$ (forward filling) was used instead of the prediction model.

For both base model and meta model a L2 regression loss function with a Huber regularizer was used, and using a separate validation set, the regularization weight alpha was optimized over the range [1.0,0.1,0.01,0.001,0.0001]. In the loss function the samples were weighted according to the formula $10+100/(1+\exp(0.025*(RealPaO_2-110)))$, to give true $PaO_2$ values close to the relevant decision boundaries for respiratory failure annotation higher weights. Model development of the $PaO_2$ estimation model did not use the held-out test set, which was not used prior to the final result preparation.

#### $F_IO_2$ estimation

For the calculation of the P/F ratio, estimates of $F_IO_2$ values were necessary for every grid point. Three situations need to be distinguished: 1) the patient is breathing ambient air, i.e. $F_IO_2$ = 21% (the ambient air oxygen fraction); 2) the patient is receiving supplemental oxygen and the corresponding $F_IO_2$ is recorded in the data 3) for patients on mechanical ventilation $F_IO_2$ is controlled by the ventilator and its value is recorded in the data. $F_IO_2$ estimation at every grid point is implemented in the following way.

a) $F_IO_2$ is forward filled from the last $F_IO_2$ measurement, if (1) it was within the last 30 minutes, and (2) the

patient was estimated to be mechanically ventilated (using the ventilation detection algorithm described later) or the ventilation mode is NIV (non-invasive ventilation).
b) Otherwise, the two supplementary oxygen variables (Supplemental $F_IO_2$ [%] and Highflow $F_IO_2$ [%]) were considered, if a measurement was available in the last 12 hours. Hereby, Supplemental $F_IO_2$ [%] takes precedence if it was available in the last 12 hours.
b) If there was no measurement in the two supplementary oxygen $F_IO_2$ variables in the last 12 hours, then an ambient air assumption was made, and $F_IO_2$ was estimated as 21 %.

#### Estimation of the P/F index

The P/F index (or ratio) at each grid point was defined as $PaO_2$ estimate / $F_IO_2$ estimate, where the $PaO_2$, $F_IO_2$ estimates at the grid point were found by the two schemas explained above. As post-processing a Nadaraya Watson kernel smoother with a bandwidth of 20 was applied to the tentative P/F indices, to yield the final estimated P/F ratios per grid time point.

#### Respiratory failure annotation

Lung function is clinically evaluated using the ratio of blood oxygen partial pressure ($PaO_2$) and fraction of inspired oxygen ($F_IO_2$) as an indicator of venous admixture, commonly referred to as P/F ratio[10]. A healthy person breathing room air is expected to have a P/F ratio of approximately 475 mmHg ($PaO_2$: ~100 mmHg, room air $F_IO_2$: 21 %). Current medical literature defines acute respiratory failure in three stages[11]:

Mild: 200 mmHg ≤ P/F ratio < 300 mmHg
Moderate: 100 mmHg ≤ P/F ratio < 200 mmHg
Severe: P/F ratio < 100 mmHg

A grid point was labeled with the 3 severity levels or ‘stable’ using a forward facing window of length 1 hour. If ⅔ of the grid points satisfied the severe criterion (<100 mmHg), it was labeled as ‘severe respiratory failure’, otherwise if ⅔ of the grid points satisfied the moderate criterion (<200 mmHg), it was labeled as ‘moderate respiratory failure’, otherwise if ⅔ of the grid points satisfied the mild criterion (<300 mmHg), it was labeled as ‘mild respiratory failure’. Otherwise the patient was labeled as ‘stable’ at the grid point. If for at least ⅔ grid points, the P/F ratio could not be estimated, the respiratory failure status of the grid point was set to ‘Unknown’. Additionally to satisfying the condition on the P/F ratio in the window, we also required that the patient was in a consistent ventilation state during the grid-points where the P/F ratio criterion was satisfied (patient is not ventilated, or patient is ventilated and PEEP is not densely available, or patient is ventilated and PEEP is densely available and satisfying PEEP>=4).

Because the labeling algorithm with a forward facing window can mis-label points on the right boundary of events as ‘not in failure’, the right edges of events were manually corrected by scanning right-wards from the tentative right edge of the event and setting the grid point to the respective severity level if the current P/F ratio actually satisfied the criterion, but was mis-labeled as not satisfying the criterion due to the forward-facing 1 hour window.

As a last step, a post processing was performed where small events (length <= 4 hours) that are sandwiched between two other events, (1) at least one of which was longer than the sandwiched event, and (2) the 2 surrounding events had the same severity label, were relabeled to match the label of the surrounding events. In this way, spuriously labeled short respiratory failure events shorter than 4 hours were deleted. Moreover small gaps between two respiratory failure events were deleted and the two events merged together.

#### Ventilation status annotation

To derive the ventilation status (binary) at each grid-point a voting algorithm was used, which was informed by prior medical knowledge. Each criterion was evaluated per grid point, and depending on the outcome, positive or negative points were assigned. Positive points correspond to a higher likelihood of ventilation at a grid point, negative points to a lower likelihood of ventilation. Finally, a cut-off on the total sum was specified, using prior medical knowledge, and by judging the correctness of endpoints visually using a time series visualization toolkit, developed for this project. Points assigned by the voting system were as follows:

a) +1 point, patient was admitted before 2009/12/06, to take into account different recording of ventilation information in the PDMS before this data
b) +2 points: In a centered 30 minute window on the grid point, at least one EtCO2 measurement of >0.5mmHg was observed, indicating active invasive or non-invasive mechanical ventilation
c) +1 point: The current estimated ventilation mode is 2 (controlled mode) or 3 (spontaneous mode)
d) -1 point: The current estimated ventilation mode is 1 (standby)
e) -2 points: The current estimated ventilation mode is 4 (NIV), 5 (High flow) or 6 (CPAP)
f) +1 point: Estimated tidal volume (TV) is >0 mL
g) +2 points: Tracheotomy indicator (vm313) or Intubation indicator (vm312) or Airway category (vm66) is 'Intubated' or 'Tracheostomy'
h) -1 point: No airway, Airway category (vm66) is 'Maske' (mask), 'Helm' (helmet), 'Mundstueck' (mouthpiece), 'Nasenmaske' (nasal interface).

If the combined score is at least 4 at a grid point the ventilation status is 'True', 'False' otherwise.

Thereafter post-processing was applied

1) We removed gaps which were likely caused by the patient leaving the ICU for procedures. The gaps were detected by missing heart rate information during the period in question (fewer than 50 % of the time points in the gap had at least one additional HR observation within 10 minutes). The patient was assumed to be ventilated during these gaps. Gaps of <15 minutes between successive ventilation episodes were removed and uninterrupted ventilation was assumed.
2) Gaps of <24 hours were closed in case a patient had a tracheotomy indicator before and after the event (normal tracheostomy weaning procedure).
3) Short ventilation events of length <45 minutes were deleted, if they did not occur at the beginning of the stay (i.e. no heart rate was recorded before the event), as these are likely spurious detections.

#### Readiness to extubate annotation

Readiness to extubate status was only annotated for time points where the patient was mechanically ventilated according to the criteria mentioned above. It was informed by medical prior knowledge and at each time-point the number of violations of commonly accepted extubation criteria was counted, to form a scoring system:

1) Ventilator mode is not 3 (spontaneous breathing), the patient cannot be extubated. A violation score of +9 is assigned. For data from prior to 2010 this criterion was not applied as the ventilator mode was sometimes incorrectly recorded in the data.
2) Current PEEP is >7: violation score of +3
3) Current pressure support is >10: violation score of +3
4) Current $F_IO_2$ is >0.4: violation score of +3
5) The rapid shallow breathing index (1000*RR/TV) is at least 105: violation score of +3
6) Current RR is at least 35 breaths/minute: violation score of +3
7) Current MV (Minute volume) is at least 10 L/min: violation score of +3
8) Current P/F ratio (as estimated using the annotation algorithm for respiratory failure) is <= 150 mmHg: violation score of +3
9) Current $PaCO_2$ is at least 50 mmHg: violation score of +3
10) Glasgow coma scale (GCS) is <= 8: violation score of +1
11) Current mean arterial pressure (MAP) is <= 60 mmHg: violation score of +1
12) Standardized dose of norepinephrine is >0.05/ug/kg/h or any dose of inotropes (epinephrine, dobutamine, milrinone, levosimendan, theophylline): violation score of +1
13) Current lactate is >=2.5 mmol/L: violation score of +1

If the sum of violation scores from the 13 criteria at any time-point is <9, the patient was assumed (tentative) to be ready for extubation. To increase the robustness of the annotation, a backward window of length 1 hour was used, and the patient was assumed to be ready for extubation if ⅔ of time-points in the last hour satisfied the criteria. The coefficients of the scoring system were obtained by fitting a model to predict extubation failure from the input variables, and then rounding the coefficients to be integers. The threshold of 9 points was determined by visually inspecting the time series annotated with integer scores, to check for clinical plausibility.

#### Extubation failure (EF) task

For the purpose of extubation failure prediction, decannulations from tracheostomy were not considered. An extubation was defined as a transition from ventilated to non-ventilated status, where the annotation algorithm for ventilation detection was used. The label for extubation failure was defined as positive, if the patient was re-intubated within the next 48 hours after the extubation event. The re-intubation was ignored if the patient was away from the ICU in the hour immediately prior to the re-intubation (detected by a HR measurement gap of ⅔ of the hour) which might indicate that the re-intubation was for procedural reasons, not for respiratory failure. If a valid reintubation occurred, the label for extubation failure was positive, otherwise negative. If the patient died within the next 48 hours after extubation, and no re-intubation occurred, the label was treated as uncertain. Sample augmentation was used for training and evaluation in the near vicinity of extubations, i.e. the prior 30 minutes before an extubation share the same label (extubation failure or no extubation failure) as the exact time-point of the extubation. In this way, the number of training samples is increased, and a clinically reasonable assumption is made that the physiological state reflecting likelihood of extubation failure does not change within a time span of 30 minutes.

#### Respiratory failure onset (RF) task

We are interested in predicting the onset of hypoxemic respiratory failure of moderate or severe level, as previously defined. The machine learning label was only defined at time-points where the patient was not already in respiratory failure (P/F ratio <200 mmHg) and the annotation was not ‘unknown’. We assigned a positive label if the patient was currently stable or in mild respiratory failure, but moderate or severe respiratory failure occurred at some point in the next 24 hours. The label was undefined if the respiratory status was ‘unknown’ at the current time-point or for the entire next 24 hours.

#### Ventilation onset ($MV_{Start}$) task

We are interested in predicting the onset of mechanical ventilation as defined by the score-based algorithm presented earlier. The machine learning label was only defined at time-points where the patient was not already mechanically ventilated. If the patient was currently not ventilated, but was mechanically ventilated at some point in the next 24 hours, the label was positive and negative otherwise. The 30 minutes just before the onset of mechanical ventilation were excluded from training and evaluation, to prevent any potential leakage of information from the future. The label was undefined if the ventilation status is ‘unknown’ for the complete next 24 hours, or at the current time-point.

#### Readiness to extubate onset ($MV_{End}$) task

We are interested in predicting if an intubated patient becomes newly ready to extubate as defined above. The machine learning label was defined at time-points where the patient was mechanically ventilated and not yet ready to be extubated. If they were currently not ready to extubate, but were ready to extubate at some point within the next 24 hours, the label was positive and negative otherwise. The label was undefined if the readiness to extubate status was ‘unknown’ for the entire next 24 hours, or at the current time-point.

#### Feature extraction

To give our model a comprehensive view of the patient state, the following feature classes were extracted from the clinical parameters available in the HiRID-II dataset.

- **Current value:** The current time grid value of the clinical parameters in the HiRID-II dataset was used directly as a feature.
- **Time since admission:** The time since admission was used as an individual feature.
- **Endpoint annotation variables:** The current estimated value of $F_IO_2$ and the current ventilation status, as computed by the scoring algorithm, were used as additional clinical variables. The current $PaO_2$ estimate was not used to avoid potential leakage of information from the future.
- **Multi-resolution summaries:** Various summary functions were computed over multiple horizons, including the last 10 hours, the last 26 hours, the last 63 hours, and the last 156 hours. These 4 horizon lengths correspond to the 20/40/60/80 percentiles of the available history across all time points in the training set. From the training set the expected number of measurements within the horizon was estimated, using the median observation interval of the parameter. If the expected number of measurements in the horizon was less than 5, the horizon was not used for feature computation. For ordinal variables, median/IQR/trend were used as the 3 summary functions. The trend was defined as the slope of a regression line fitted over the values in the horizon. For binary variables, the mean was used as the only summary function. Note, for binary

variables, the mean can be also interpreted as the proportion of the horizon in which a certain condition was true. For categorical variables, the mode was used as the only summary function. All 4 horizons were computed only for important variables which were determined using a preliminary variable importance selection analysis. The important variables are listed in Supplemental Table 3. For other variables, only the shortest of the 4 horizons, for which the expected number of measurements exceeded 5, was used.

- **Measurement intensity:** The time to last real measurement was computed as a feature. If there was no such measurement, this feature was set to a very large value. The measurement density was computed over the same multi-resolution horizons as in the previous feature category. The measurement density was defined as the number of observations in the horizon divided by the horizon length.
- **Instability history:** If applicable for a variable, up to 3 severity levels were annotated using prior medical knowledge. The fraction of time spent in each severity state over the last 8 hours as well as over the entire stay up to the current time point was extracted. This schema was used only for a subset of variables, which were among the important variables selected in a preliminary variable selection step. The severity levels and variables used for this feature class are listed in Supplemental Table 4.
- **Static variables:** Static variables are constant for all time points of the patient's time series and are finally concatenated to the feature vector. As static variables, the patient age, APACHE patient group, gender, Emergency admission status, Surgical admission status, and height were used.

#### Variable selection

Variable selection was performed in a 2-step process using only the development set, and not the held-out test set.

a) The 20 most important variables in terms of SHAP value magnitude were pre-selected on the validation set for the 4 tasks (Respiratory failure, Extubation failure, Ventilation onset, Readiness to extubate) separately. The 'SHAP importance' of a feature was defined as the mean (over the 5 temporal splits) of the mean absolute SHAP value on predictions in the validation set for that variable. The 'SHAP importance' of a variable was defined as the maximum of SHAP importance over the features derived from the clinical variable. In this way 20 variables were extracted per task. The union of the variables selected for the 4 tasks formed the initial set of 'important variables', which consisted of 31 variables, and are listed in Supplemental Table 3.
b) For the initial set of important variables, more complex features were computed, according to the description in the section on feature extraction above.
c) For the 2 main tasks presented in the work, Respiratory failure (RMS-RF) and Extubation failure (RMS-EF), variables were greedily forward selected from the final set of complex features on 31 variables. In each step the variable was chosen, which yielded the highest time-point based AUPRC on the validation set, among the candidate variables to be added. The output of this procedure, which was run 5 times per temporal split, was a forward trace of 31 variables, ranked by importance. The final importance of a variable was defined as the mean reciprocal rank over the 5 splits, yielding a ranked list of 31 variables. The RMS-EF model used the top 20 variables, which included both medication and non-medication variables, and the RMS-RF model used the non-medication variables among the top 20 variables, which yielded a final set of 15 variables. For ventilation onset/readiness to extubate prediction models, the union of the variables used for the respiratory failure and extubation failure models was used. The variables used by each model are listed in Supplemental Table 3.

#### Model training

The generated features for the variable sets of the RMS-RF (15 variables) and RMS-EF (20 variables), RMS-$MV_{Start}$/RMS-$MV_{End}$ (26 variables) predictors were passed to 4 gradient-boosted decision tree ensembles implemented in LightGBM (https://lightgbm.readthedocs.io/en/latest/pythonapi/lightgbm.LGBMClassifier.html), with one separate model per task. As LightGBM is robust to missing data and different feature scales, data was not imputed or standard scaled prior to training. Trees were added to the ensemble until performance did not improve for 50 epochs in the validation set, early stopping the training process. As a criterion guiding the early stopping the time-point based AUPRC on the validation set was used. Hyperparameters were optimized on the validation set, using the time-point based AUPRC as a criterion. Hyperparameters were fixed for RMS-RF/RMS-$MV_{Start}$/RMS-$MV_{End}$, as experiments showed that early stopping was enough to find a configuration close to optimal. For RMS-EF, which had a small number of training set samples, a hyperparameter grid with 20 points was used to select the optimal model. The parameters of the LightGBM model and the hyperparameter selection grid for RMS-EF are listed in Supplemental table 5. Prediction scores were generated for patients in the test set, using a separate LightGBM model for each of the 4 tasks. To allow more flexible evaluation, for resource planning and joint task analysis using t-SNE, predictions were generated at all time-points of test set patients, even when the ground-truth label was undefined, i.e. for RMS-RF, while the patients were already experiencing respiratory failure.

#### Resource planning

We developed a resource planning model for mechanical ventilation that predicts how many patients will require mechanical ventilation within certain time-windows in the short-term future. This includes the start of mechanical ventilation for patients who are already admitted to ICU but not yet ventilated and additional non-elective patient admissions requiring mechanical ventilation. As the original dates were removed during anonymization for HIRID-II, we used an additionally provided dataset with the admission dates of the ICU patients in order to reconstruct the number of patients within the ICU and the ventilator resource use. To predict number of ICU patients who will require ventilation in the near future, we used the outputs from the machine learning models trained for predicting the four respiratory system related tasks (respiratory failure, mechanical ventilation need, readiness to extubate and the extubation failure) for individual patients, as well as ICU-level information as features for the LightGBM model. The ICU-level information included the hour of the day, weekday, and number of patients who were admitted and required mechanical ventilation in the past hour. To predict newly admitted emergency patients who will require mechanical ventilation, we trained a LightGBM model using only the ICU-level information.

#### Model calibration

For the evaluation of model calibration, the prediction scores of time-points in the test set where the label was defined were gathered. The scores were binned between the minimum and maximum prediction scores observed in the test set, using a bin size of 0.05. The actual observed risk (proportion of true labels for time-points in the bin) was then computed per bin and plotted against the bin location. As an evaluation metric of calibration the Brier score [12] was used. Models showed sufficient calibration using the raw scores, so re-calibration using isotonic regression was not needed.

#### Prevalence correction for external validation

As the prevalence of positive events was different between the HiRID-II and the UMCdb test sets, we corrected the precision-recall curves for the performance on the test set of UMCdb such that the corrected prevalence matches with that in the HiRID-II test set by downscaling the false alarm number using the scaling factor s = (1/prev(HiRID-II)-1)/(1/prev(UMCDB)-1), as used by Hyland et al. [5]

#### Extubation-based evaluation (extubation failure)

Extubation failure was assessed using recall (percentage of extubation failures which were correctly predicted), and precision (percentage of extubation failure predictions which are correct, i.e. re-intubation occurs in the next 48h), yielding a Precision-Recall curve, as well as recall/false positive rate, which defined the ROC curve.

#### Event-based evaluation (respiratory failure)

We used the same event-based evaluation scheme used by Hyland et al.[5], which measures the fraction of correctly predicted respiratory-failure events (recall) and the fraction of false-alarms (1-precision).

#### External validation + prevalence correction

To allow external validation, the most important parameters for training predictive models in HiRID-II were matched to variables in the Amsterdam UMCdb dataset[1]. To enable endpoint annotation at a similar granularity as in HiRID-II, a subset of patients with high time resolution for respiratory parameters (n=6,698 patients) was included. A dataset was then prepared by applying the same endpoint annotation and feature extraction pipeline as for HiRID-II. Because admission times are not exactly available in UMCdb, data were randomly divided by patient into a fixed test set containing 1,674 patients, and a development dataset of 5,024 patients. To retrieve variation estimates of performance, the development dataset was partitioned five times into training and validation sets. For external validation, we applied the models trained on HiRID-II dataset to the UMCdb dataset, and used prevalence correction to re-scale the false positive count, as described in the section on ‘Prevalence correction’.

#### Sub-cohort / Fairness analyses

Patients grouping. We grouped patients by demographic characteristics (sex, age) and clinical characteristics (APACHE II/IV admission group). For binary grouping, we compared one group versus the other, while for multi-categorical grouping we compared patients belonging to a group to all the other patients.

Patients bootstrapping. Due to the small number of patients composing certain cohorts of patients, we relied on bootstrapping. We created 100 bootstrap samples of the patients from the test set (i.e. we sample randomly with replacement from the patients composing the test set). We then computed the different performance metrics for each bootstrap sample and for each of our patient cohorts. Having several bootstrap samples to perform the analysis on, allowed us to better understand the variability of the patients within each cohort and to present these in the result section.

Statistical testing. To compare groups of patients we assumed that the samples of one cohort of patients and the other cohort are independent. However, we do not assume normality of the distribution. To compare the distribution of our metrics across different cohorts, we used the Mann-Whitney U test at a significance level of 0.1%. Since for each grouping we performed multiple tests, we corrected the p-value with a Bonferroni correction. We tested whether patients from a certain group are significantly worse off (according to fairness of prediction performance) compared to patients not belonging to this group. On the result plots, we mark groups that are significantly worse off in terms of prediction performance with a star.

#### Metrics used

For the respiratory failure task, we computed the precision at:

- 80% event-based recall for each cohort (the threshold will thus be different for each cohort)
- 90% event-based recall for each cohort (the threshold will thus be different for each cohort)

For the extubation failure task, we computed the precision at:

- 80% recall for each cohort (the threshold will thus be different for each cohort)
- 20% recall for each cohort (the threshold will thus be different for each cohort)

Finally, for both tasks, we also computed the corrected event-based AUPRC.

#### Model inspection using SHAP values

SHAP values for the positive class were extracted using a 'SHAP tree explainer' built for LightGBM ensembles (https://shap-lrjball.readthedocs.io/en/latest/generated/shap.TreeExplainer.html), in the validation set and test set. In the validation set the mean absolute SHAP values of each feature were used to create an initial set of important variables for variable selection (refer to the section on 'Variable selection' above). In the test set the signed SHAP values (Interpretation: Large SHAP value means the feature value contributes to an increase of the prediction score) were used to interpret the model's prediction, i.e. by plotting them against the feature value at the time-point when the prediction is made.

#### Joint task analysis using t-SNE

For the joint task analysis the test set predictions for the 4 tasks (RMS-RF/RMS-EF/RMS-$MV_{Start}$/RMS-$MV_{End}$) were used. In principle, predictions were available at all time points in the patient stay for all tasks, irrespective of whether the label of the task was defined at the time-point. For computing the t-SNE embedding, only the current value features of 16 clinical parameters were used, which correspond to the union of the top 10 important variables for the RMS-RF/RMS-EF models. As t-SNE requires dense input without missing values, the 'dense imputation' data (refer to the section on Imputation for its definition), was used as an input for t-SNE. Prior to fitting of the t-SNE map the data was standard scaled such that each dimension had mean 0 and standard deviation 1, such that all variables have equal importance in the t-SNE input space. A random subsample of 150,000 time-points in the test set was drawn to allow fast fitting of the embedding algorithm. The t-SNE map was computed once, independent of task, as it only depends on the clinical parameters but not on the prediction scores of the 4 models. For fitting t-SNE, the implementation available in the Python package scikit-learn (https://scikit-learn.org/stable/modules/generated/sklearn.manifold.TSNE.html) was used with default parameters, and target dimension 2. In the t-SNE plots, only hexes with at least 30 assigned time-points were displayed, ignoring very rarely used parts of the embedding space.

#### Statistical methods

In result plots the solid curves refer to the mean of the performances obtained in the five experimental replicates, corresponding to the five temporal splits, as described in the section on data splits. Light shaded regions refer to the standard deviation of performances obtained in the five experiment replicates.

#### Data availability

More information on HiRID is available on hirid.intensive care.ai. The newly curated data set HiRID-II will be released on Physionet in the near future. The computer code used in this research is available at www.github.com/ratschlab/RMS under an open-source license.

### References


1. Thoral, P. J. *et al.* Sharing ICU Patient Data Responsibly Under the Society of Critical Care Medicine/European Society of Intensive Care Medicine Joint Data Science Collaboration: The Amsterdam University Medical Centers Database (AmsterdamUMCdb) Example. *Crit. Care Med.* **49**, e563–e577 (2021).
2. Faltys, M., Zimmermann, M., Lyu, X., Hüser, M. & Hyland, S. HiRID, a high time-resolution ICU dataset (version 1.1. 1). *Physio. Net* (2021).
3. Johnson, A. E. W. *et al.* MIMIC-III, a freely accessible critical care database. *Sci Data* **3**, 160035 (2016).
4. Amsterdam Medical Data Science. AmsterdamUMCdb website and documentation. https://amsterdammedicaldatascience.nl/#amsterdamumcdb.
5. Hyland, S. L. *et al.* Early prediction of circulatory failure in the intensive care unit using machine learning. *Nat. Med.* **26**, 364–373 (2020).
6. Ke, G. *et al.* LightGBM: A Highly Efficient Gradient Boosting Decision Tree. in *Advances in Neural Information Processing Systems 30* 3146–3154 (2017).
7. Brown, S. M. *et al.* Nonlinear Imputation of PaO2/FIO2 From SpO2/FIO2 Among Mechanically Ventilated Patients in the ICU: A Prospective, Observational Study. *Crit. Care Med.* **45**, 1317–1324 (2017).
8. Ellis, R. K. Determination of PO2 from saturation. *J. Appl. Physiol.* **67**, 902 (1989).
9. Severinghaus, J. W. Simple, accurate equations for human blood O2 dissociation computations. *J. Appl. Physiol.* **46**, 599–602 (1979).
10. Bernard, G. R. *et al.* The American-European Consensus Conference on ARDS. Definitions, mechanisms, relevant outcomes, and clinical trial coordination. *Am. J. Respir. Crit. Care Med.* **149**, 818–824 (1994).
11. ARDS Definition Task Force *et al.* Acute respiratory distress syndrome: the Berlin Definition. *JAMA* **307**, 2526–2533 (2012).
12. Brier, G. W. Verification of forecasts expressed in terms of probability. *Mon. Weather Rev.* **78**, 1–3 (1950).
